# Evolution of long-term vaccine induced and hybrid immunity in healthcare workers after different COVID-19 vaccination regimens: a longitudinal observational cohort study

**DOI:** 10.1101/2022.06.06.22275865

**Authors:** Shona C. Moore, Barbara Kronsteiner, Stephanie Longet, Sandra Adele, Alexandra S. Deeks, Chang Liu, Wanwisa Dejnirattisai, Laura Silva Reyes, Naomi Meardon, Sian Faustini, Saly Al-Taei, Tom Tipton, Luisa M Hering, Adrienn Angyal, Rebecca Brown, Alexander R Nicols, Susan L Dobson, Piyada Supasa, Aekkachai Tuekprakhon, Andrew Cross, Jessica K Tyerman, Hailey Hornsby, Irina Grouneva, Megan Plowright, Peijun Zhang, Thomas A.H. Newman, Jeremy M. Nell, Priyanka Abraham, Mohammad Ali, Tom Malone, Isabel Neale, Eloise Phillips, Joseph D. Wilson, Sam M. Murray, Martha Zewdie, Adrian Shields, Emily C. Horner, Lucy H. Booth, Lizzie Stafford, Sagida Bibi, Daniel G. Wootton, Alexander J. Mentzer, Christopher P. Conlon, Katie Jeffery, Philippa C. Matthews, Andrew J. Pollard, Anthony Brown, Sarah L. Rowland-Jones, Juthathip Mongkolsapaya, Rebecca P. Payne, Christina Dold, Teresa Lambe, James E.D. Thaventhiran, Gavin Screaton, Eleanor Barnes, Susan Hopkins, Victoria Hall, Christopher JA Duncan, Alex Richter, Miles Carroll, Thushan I. de Silva, Paul Klenerman, Susanna Dunachie, Lance Turtle, the PITCH Consortium

## Abstract

Both infection and vaccination, alone or in combination, generate antibody and T cell responses against SARS-CoV-2. However, the maintenance of such responses – and hence protection from disease – requires careful characterisation. In a large prospective study of UK healthcare workers (Protective immunity from T cells in Healthcare workers (PITCH), within the larger SARS-CoV-2 immunity & reinfection evaluation (SIREN) study) we previously observed that prior infection impacted strongly on subsequent cellular and humoral immunity induced after long and short dosing intervals of BNT162b2 (Pfizer/BioNTech) vaccination. Here, we report longer follow up of 684 HCWs in this cohort over 6-9 months following two doses of BNT162b2 or AZD1222 (Oxford/AstraZeneca) vaccination and up to 6 months following a subsequent mRNA booster vaccination. We make three observations: Firstly, the dynamics of humoral and cellular responses differ; binding and neutralising antibodies declined whereas T and memory B cell responses were maintained after the second vaccine dose. Secondly, vaccine boosting restored IgG levels, broadened neutralising activity against variants of concern including omicron BA.1, BA.2 and BA.5, and boosted T cell responses above the 6 month level post dose 2. Thirdly, prior infection maintained its impact driving larger as well as broader T cell responses compared with never-infected people – a feature maintained until 6 months after the third dose. In conclusion, broadly cross-reactive T cell responses are well maintained over time – especially in those with combined vaccine and infection-induced immunity (“hybrid” immunity) – and may contribute to continued protection against severe disease.

## INTRODUCTION

As vaccines have been deployed to tackle the SARS-CoV-2 pandemic, crucial questions have emerged regarding long-term maintenance of protective immunity against disease. The appearance of viral variants leading to successive waves of infection has clearly shown the limits of vaccine protection against infection (UK Health Security Agency, 2022). Despite this, vaccine protection against severe disease has been well maintained across the recent delta (Tartof et al., 2021) and omicron BA.1 (Andrews et al., 2022) waves. To understand the underlying immune responses that determine these population-level observations, large-scale studies of individuals with high exposure to SARS-CoV-2, such as health care workers (HCWs), can provide valuable insights as has been demonstrated by the SARS-CoV-2 immunity & reinfection evaluation (SIREN) study in the UK (Hall et al., 2022; Hall et al., 2021a; Hall et al., 2021b). Protective Immunity from T Cells in Healthcare workers (PITCH), a study aligned closely with SIREN, is focused on the longitudinal analysis of antiviral T and B cell responses after infection and/or vaccination with BNT162b2 (Pfizer/BioNTech) or AZD1222 (Oxford/AstraZeneca). PITCH has already provided data indicating that the extended interval vaccine regimen for BNT162b2 mRNA vaccine deployed in the UK was associated with enhanced antibody and CD4^+^ T cell helper responses (Payne et al., 2021a). All immune responses were strongly enhanced by prior SARS-CoV-2 infection.

The long-term impacts of prior exposure, vaccine regimen and vaccine type have not been fully defined, especially at the level of T cell responses. Characterising the response to vaccines and infections in healthy people is essential to determine future vaccination policies, while identification of vulnerable non-responders can inform additional interventions such as extra booster doses of vaccine and/or monoclonal antibody therapies. Correlations with protection from infection at a population level have been observed for binding (Earle et al., 2021; Gilbert et al., 2022) and neutralising antibodies (Addetia et al.; Feng et al., 2021; Gilbert et al., 2022; Khoury et al., 2021; Moore et al., 2021). The role of other, non-neutralising antibody functions, such as antibody-dependent NK cell activity, antibody-dependent phagocytosis or complement deposition, requires further investigation (Ewer et al., 2021; Kaplonek et al., 2022a; Tomic et al., 2022). However, monitoring of SARS-CoV-2 specific T cell immunity is also essential, as T cell defence is potentially a key explanation for lower case hospitalisation and mortality for the omicron variant compared with earlier variants (Nyberg et al., 2022), despite omicron’s high mortality in unvaccinated populations (Mefsin et al., 2022). T cells are a cornerstone of antiviral defence, orchestrating the immune response including cytotoxic activity against virally infected cells and optimising production of antibodies from B cells (Sette and Crotty, 2021). Macaque (McMahan et al., 2021) and human (Kedzierska and Thomas, 2022; Molodtsov et al., 2022; Rydyznski Moderbacher et al., 2020) studies support this key role for T cells in protection against the severe effects of SARS-CoV-2 infection, potentially alongside functional antibody properties beyond neutralisation (Bartsch Yannic et al.; Kaplonek et al., 2022b). In some cases, cross-reactive T cells are associated with protection against infection in exposed seronegative groups (Kundu et al., 2022). There is also evidence of SARS-CoV-2 specific cell responses in highly exposed HCW without seroconversion (Ogbe et al., 2021), and expansion of pre-existing RNA-polymerase-specific T cells in seronegative SARS-CoV-2 infection (Swadling et al., 2022).

There is a body of emerging data on the waning of antibody responses, especially after the shorter dose interval regimen for BNT162b2 (Goldberg et al., 2021; Naaber et al., 2021). Waning of antibody is associated with loss of protection against infection (Hall et al., 2022; Wei et al., 2022b), whereas protection against severe disease is relatively well maintained (Andrews et al., 2022; Carazo et al., 2022; Lin et al., 2022; Rosenberg et al., 2021; Tartof et al., 2021; UK Health Security Agency, 2022). T cell responses to spike protein post vaccination do not correlate strongly with binding or neutralising antibody responses (Payne et al., 2021a). Importantly, whilst antibodies generated in response to vaccination neutralise omicron much less well than the ancestral strain (Dejnirattisai et al., 2022; Schmidt et al., 2021), the T cell response to SARS-CoV-2 is minimally impacted by mutations in the alpha, beta, gamma and delta variants of concern (Payne et al., 2021a; Skelly et al., 2021), and 75-85% preserved against the omicron BA.1 variant (De Marco et al., 2022; Gao et al., 2022; GeurtsvanKessel Corine et al., 2022; Keeton et al., 2022; Liu et al., 2022; Madelon et al., 2022; Tarke et al., 2022). Given that at this point in the pandemic, public health decisions are increasingly being made around limiting severe disease rather than preventing milder infections in the community, having robust data at scale that indicates the trajectory of the T cell responses after different vaccine regimens is of increasing value. The impact of subsequent vaccine dosing on T and B cell responses is additionally a key focus in such decision making.

We previously observed higher anti-spike binding, higher neutralising antibody responses and lower spike-specific T cell magnitude but increased IL2 production one month after second dose when BNT162b2 was delivered with a longer dosing interval (median 10 weeks) compared to the licensed shorter (3-4 weeks) interval (Payne et al., 2021a). This pattern was reproduced in an elderly population (Parry et al., 2022), and the antibody findings have been confirmed in the larger SIREN cohort (Otter et al., 2022). Evidence of improved vaccine effectiveness with a longer dose interval was reported in a study of two Canadian provinces (Skowronski et al., 2022).

In the study presented here, our objective was to explore the characteristics of adaptive and humoral immunity following two and three vaccine doses, to consider the longer-term impacts of regimen variation, vaccine type (including the Oxford-AZ ChadOx1-based vaccine) and infection over time. We observed the long-term impact of prior infection even after two doses of vaccine, which is consistent with protection documented in SIREN (Hall et al., 2022). We saw no decline in T cell responses over time regardless of vaccine regimen — this contrasts with waning of both binding and neutralising antibody (NAb) titres, which remained strongest and broadest in the long interval BNT162b2 group. The third dose of vaccine boosted binding antibody responses such that differences seen between vaccine regimens after only two doses were reduced as were differences associated with prior infection. Overall, the data indicate a stable pool of T cell memory is induced and maintained across vaccine types/regimens, consistent with the sustained impact of vaccination with or without prior infection in protection against severe disease.

## RESULTS

### Participants vaccinated with a primary course and a booster dose of COVID vaccine

We studied 684 participants who had been vaccinated with a primary course of COVID-19 vaccine between 9th December 2020 and 23rd May 2021 (Table 1 and Figure 1). In total, 592 participants received a primary course of BNT162b2 vaccine (Pfizer), of whom 84 participants received the second dose of BNT162b2 vaccine after a short (3-5 week, median 24 days) interval, and 508 participants received the second dose of BNT162b2 vaccine after an extended (6-17 week, median 71 days) interval (Payne et al., 2021a). 92 participants received a primary course of AZD1222 vaccine administered with an interval of 7-23 weeks (median 74 days). The median age of all participants was 43 (range 22-77), and 73.8% of participants were female, reflecting the demographic of healthcare workers in the UK and consistent with our previous reports, and the wider SIREN cohort.

**Figure 1.**
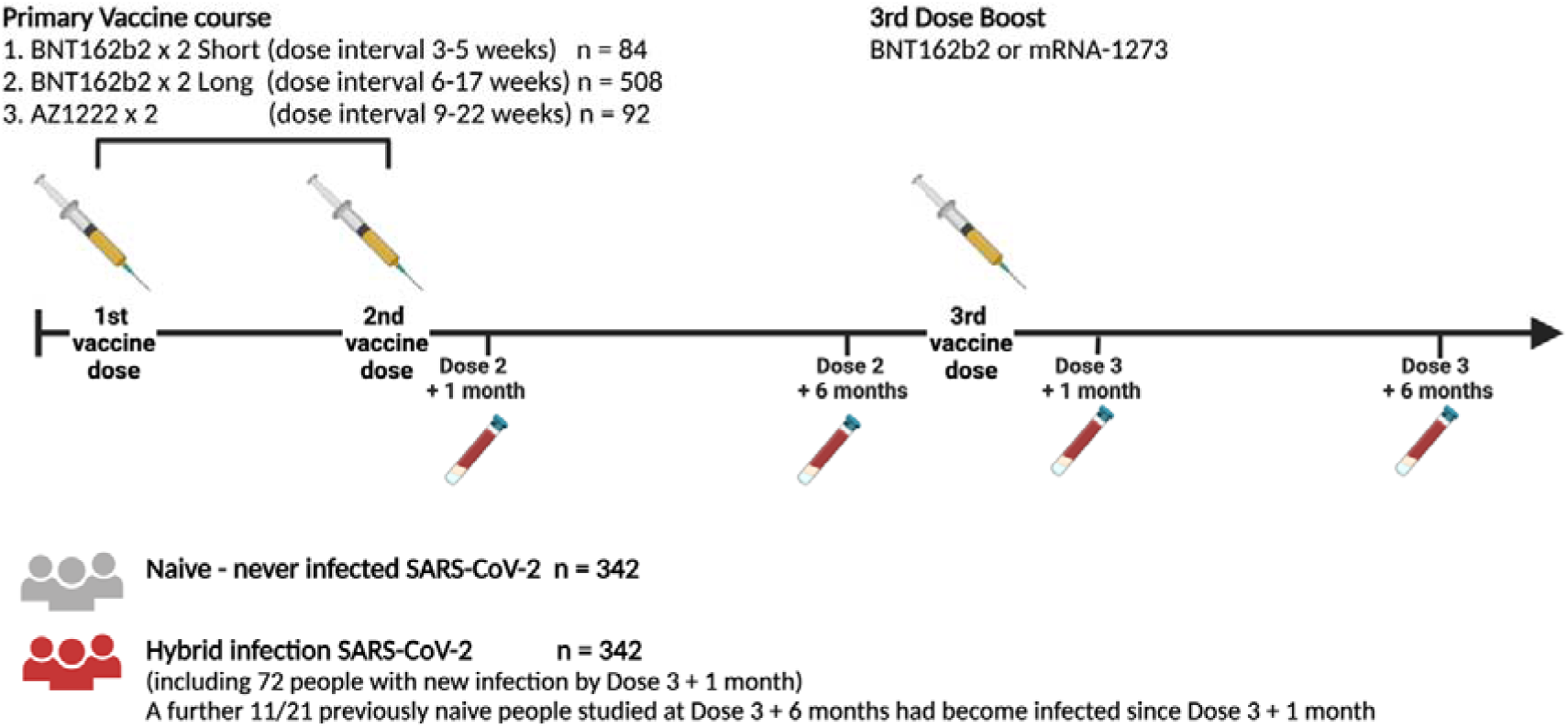
Study Design. Schematic representation of vaccination and phlebotomy time points. Figure created using Biorender.

**Table 1.**
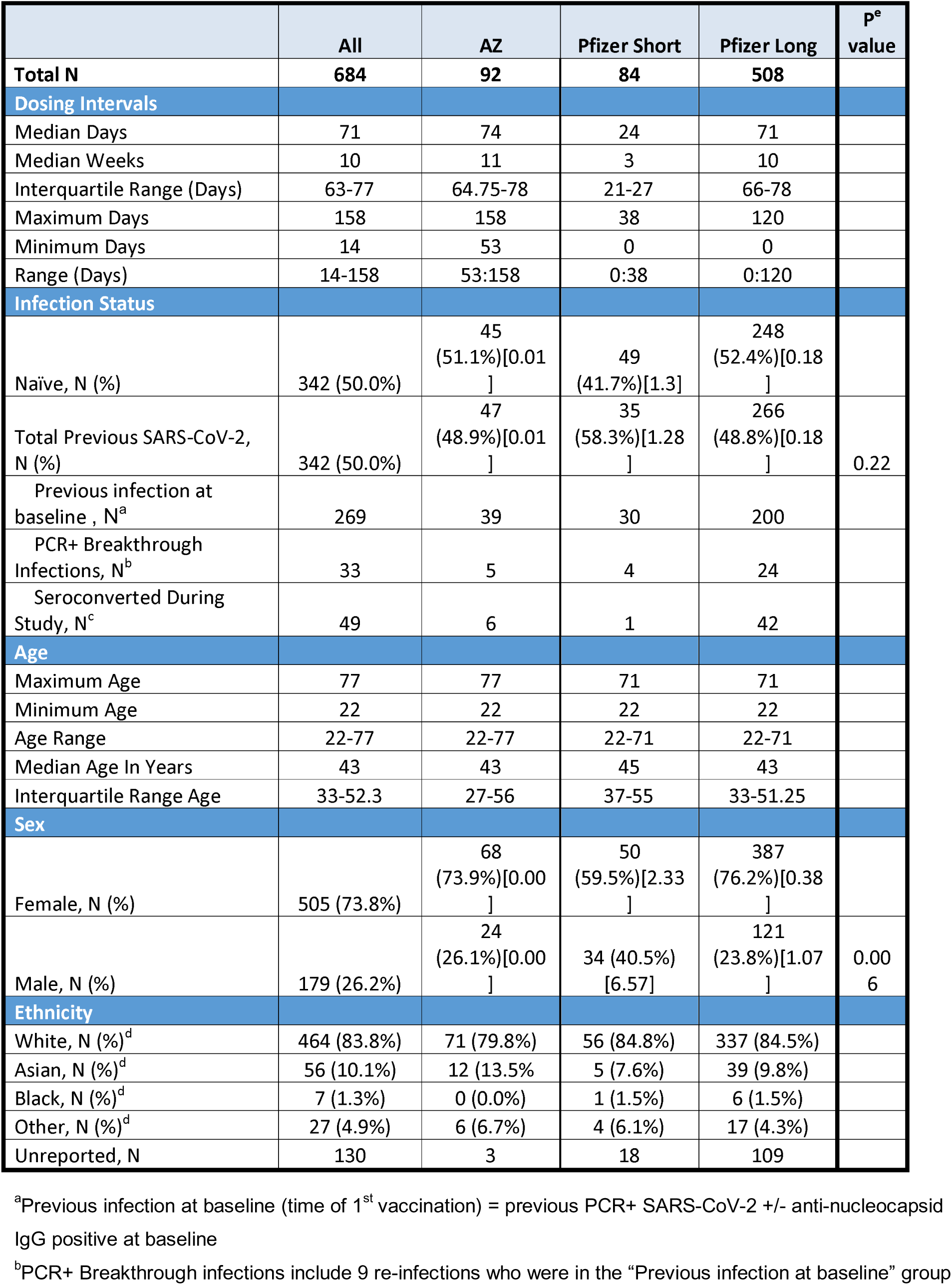
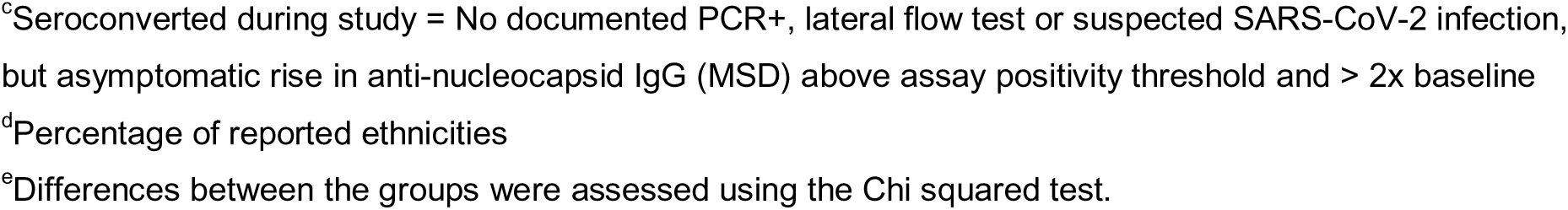
Demographic characteristics of participants in the study

|  | All | AZ | Pfizer Short | Pfizer Long | P<sup>e</sup> value |
| --- | --- | --- | --- | --- | --- |
| **Total N** | **684** | **92** | **84** | **508** |  |
| **Dosing Intervals** |  |  |  |  |  |
| Median Days | 71 | 74 | 24 | 71 |  |
| Median Weeks | 10 | 11 | 3 | 10 |  |
| Interquartile Range (Days) | 63-77 | 64.75-78 | 21-27 | 66-78 |  |
| Maximum Days | 158 | 158 | 38 | 120 |  |
| Minimum Days | 14 | 53 | 0 | 0 |  |
| Range (Days) | 14-158 | 53:158 | 0:38 | 0:120 |  |
| **Infection Status** |  |  |  |  |  |
| Naïve, N (%) | 342 (50.0%) | 45<br>(51.1%)[0.01<br>] | 49<br>(41.7%)[1.3] | 248<br>(52.4%)[0.18<br>] |  |
| Total Previous SARS-CoV-2, N (%) | 342 (50.0%) | 47<br>(48.9%)[0.01<br>] | 35<br>(58.3%)[1.28<br>] | 266<br>(48.8%)[0.18<br>] | 0.22 |
| Previous infection at baseline, N<sup>a</sup> | 269 | 39 | 30 | 200 |  |
| PCR+ Breakthrough Infections, N<sup>b</sup> | 33 | 5 | 4 | 24 |  |
| Seroconverted During Study, N<sup>c</sup> | 49 | 6 | 1 | 42 |  |
| **Age** |  |  |  |  |  |
| Maximum Age | 77 | 77 | 71 | 71 |  |
| Minimum Age | 22 | 22 | 22 | 22 |  |
| Age Range | 22-77 | 22-77 | 22-71 | 22-71 |  |
| Median Age In Years | 43 | 43 | 45 | 43 |  |
| Interquartile Range Age | 33-52.3 | 27-56 | 37-55 | 33-51.25 |  |
| **Sex** |  |  |  |  |  |
| Female, N (%) | 505 (73.8%) | 68<br>(73.9%)[0.00<br>] | 50<br>(59.5%)[2.33<br>] | 387<br>(76.2%)[0.38<br>] |  |
| Male, N (%) | 179 (26.2%) | 24<br>(26.1%)[0.00<br>] | 34 (40.5%)<br>[6.57] | 121<br>(23.8%)[1.07<br>] | 0.00<br>6 |
| **Ethnicity** |  |  |  |  |  |
| White, N (%)<sup>d</sup> | 464 (83.8%) | 71 (79.8%) | 56 (84.8%) | 337 (84.5%) |  |
| Asian, N (%)<sup>d</sup> | 56 (10.1%) | 12 (13.5%) | 5 (7.6%) | 39 (9.8%) |  |
| Black, N (%)<sup>d</sup> | 7 (1.3%) | 0 (0.0%) | 1 (1.5%) | 6 (1.5%) |  |
| Other, N (%)<sup>d</sup> | 27 (4.9%) | 6 (6.7%) | 4 (6.1%) | 17 (4.3%) |  |
| Unreported, N | 130 | 3 | 18 | 109 |  |

<sup>a</sup>Previous infection at baseline (time of 1<sup>st</sup> vaccination) = previous PCR+ SARS-CoV-2 +/- anti-nucleocapsid IgG positive at baseline
<sup>b</sup>PCR+ Breakthrough infections include 9 re-infections who were in the "Previous infection at baseline" group
<sup>c</sup>Seroconverted during study = No documented PCR+, lateral flow test or suspected SARS-CoV-2 infection, but asymptomatic rise in anti-nucleocapsid IgG (MSD) above assay positivity threshold and > 2x baseline
<sup>d</sup>Percentage of reported ethnicities
<sup>e</sup>Differences between the groups were assessed using the Chi squared test.

### Symptomatic infection and asymptomatic anti-nucleocapsid (N) seroconversion were common during the study period

During follow up of this cohort (May 2021 to March 2022), some participants became infected during the SARS-CoV-2 waves of delta and omicron BA.1 or BA.2 (Table 1). 33 participants developed symptomatic COVID-19 confirmed by positive SARS-CoV-2 PCR assay. A further 49 participants had evidence of asymptomatic infection between one and 6 months after the second vaccine, reflected by SARS-CoV-2 N antibody seroconversion detected in the 6 month samples. After accounting for those infections, half the cohort (342 participants) met the definition of infection-naïve at the time of the third vaccination. In addition, 11 participants of 21 followed up to 6 months post third dose became infected with omicron variants.

We measured T cell responses 6 months after the second vaccine dose and found that participants infected between one and 6-months after the second dose had similar T cell responses to those infected prior to their first vaccine dose (Figure S2). Spike IgG, measured by MesoScale Discovery (MSD), was lower in those infected during the study compared with those infected before vaccination, but was higher than infection naïve participants. Therefore, in this report, participants with natural infection at any point were analysed together as a “hybrid immunity” group, regardless of when the infection occurred in relation to vaccine doses.

### Six months post second vaccination, T cell IFNγ ELISpot responses are greatest following BNT162b2 short dose interval at six months and are augmented in participants with hybrid immunity

In infection-naïve participants, at 6 months post vaccine dose 2, there was no significant difference in the T cell response by IFNγ ELISpot assay between the three primary vaccine groups, although there was a trend towards higher T cell responses in those who received BNT162b2 vaccine with a short interval (median 3 weeks) than those groups who were vaccinated with a BNT162b2 long interval (median 10 weeks), or the group vaccinated with AZD1222 (Figure 2A). This difference was significant for the BNT162b2 short and long interval groups one month after the second dose (Payne et al., 2021a). Spike-specific T cell responses 6 months after the second vaccination were considerably greater in all groups (105 SFU/10^6^ PBMC, IQR 48 – 240) than the historical median responses we observed using the same assay in this cohort pre-vaccination in 2020 (Tomic et al., 2022) 6 months after wave 1 infection (44 SFU/10^6^ PBMC, IQR 1-107).

**Figure 2.**
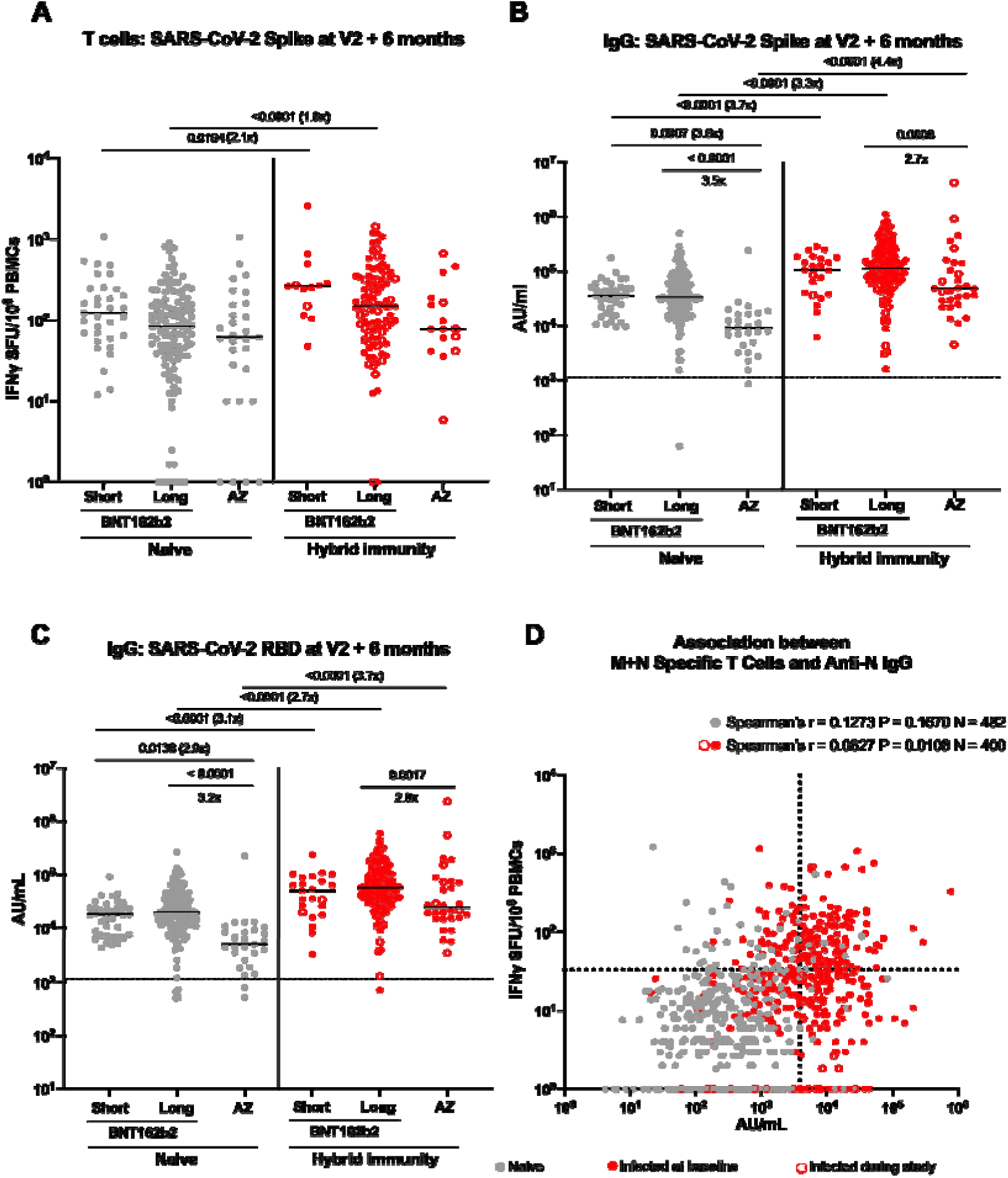
Comparison of T cell and IgG responses six months after the second dose of vaccine according to vaccine regime and infection status. (A) Comparison of IFNγ ELISpot responses to spike (S, ancestral strain) from cryopreserved peripheral blood mononuclear cells (PBMCs) in naïve (grey circles) participants 6 months after 2 doses of BNT162b2 (Pfizer-BioNTech) delivered with a short dosing interval (“Short”, 3-5 weeks, n=33), or a long dosing interval (“Long”, 6-17 weeks, n=116), or 6 months after 2 doses of AZD1222 (AstraZeneca) vaccine (“AZ”, n=29); or previously infected (closed red circles infected at baseline, open red circles infected during study) BNT162b2 short (n=13), previously infected BNT162b2 long (n=94), and AZ (n=16) vaccinated individuals. (B) Effect of vaccine regimen and infection status on SARS-CoV-2 S-specific IgG responses in naïve short (n=38), long (n=170), and AZ (n=39); and previously infected short (n=18), long (n=99), and AZ (n=28) vaccinated individuals. (C) Effect of vaccine regime and infection status on SARS-CoV-2 RBD-specific IgG responses in naïve short (n=38), long (n=169), and AZ (n=37); and previously infected short (n=18), long (n=99), and AZ (n=28) vaccinated individuals. (D) Association of membrane (M) and nucleocapsid (N) protein specific T cell and SARS-CoV-2 N-specific IgG responses in participants 6 months after second dose, and 28 days after third dose (hence participants can have >1 value), by infection status. ELISpot values are expressed as spot forming units per million (SFU/10^6^) PBMCs. Data displayed are responses to peptide pools representing the sum of S1 and S2 units of S (ancestral strain). IgG responses were measured in serum 6 months after the second dose using multiplexed MSD immunoassays and are shown in arbitrary units (AU)/mL. Bars represent the median. Vaccine regimes and infection status were compared with Kruskal-Wallis and Dunn’s multiple comparisons test (A-C) and Spearman’s tests (D), with 2-tailed p-values shown above linking lines. Where p-values are absent, comparison was not statistically significant (p>0.05). Dashed lines in (D) represent thresholds for a positive response: SARS-CoV-2 N IgG - based on the mean concentrations measured in 103 pre-pandemic sera + 3 Standard Deviations (3874 AU/ml); SARS-CoV-2 M & N IFNγ ELISpot assay – mean + 2 Standard Deviations of the DMSO wells across all experiments in the study (33 SFU/10^6^).

For anti-spike binding antibody responses, levels were higher for BNT162b2 recipients than AZD1222 recipients irrespective of the dosing interval (Figure 2B). A similar pattern was apparent for RBD antibody (Figure 2C). As was observed at one month post second dose, T cell and antibody responses were greater in magnitude in those who were previously infected at any point before the 6-month post second dose sample was collected (Figure 2A-C). T cell responses against M and N were, as expected, higher in those with hybrid immunity, and correlated with N antibody levels (Figure 2D).

### After a booster (third) vaccine, IFNγ ELISpot T cell responses are equivalent in all groups irrespective of primary vaccine regimen

Over the 6-month period following the second vaccine dose, T cell IFNγ responses were well maintained, with a modest fall which did not reach statistical significance, and, overall, were boosted significantly after the third dose in both naïve and hybrid immune participants (Figure 3A). This apparent boost was accounted for by the largest group, the BNT162b2 long interval group (Supplementary Figure S3A, D and G), whilst the other, smaller, groups did not achieve statistical significance. These responses were well maintained for 6 months following the third dose, with no significant change in T cell response between 1 and 6 months post third dose, although fewer hybrid immune participants were tested at this timepoint (Figure 3A). One month after the third vaccine dose, participants receiving all three vaccine regimens had equivalent T cell IFNγ responses (Figure 3D). The post dose 3 boosting effect did not generate T cell responses any higher than those measured 28 days post dose 2, but responses were higher than those measured 6 months post dose 2 value. Thus, all groups derived a detectable benefit on the T cell response from the third vaccine dose.

**Figure 3.**
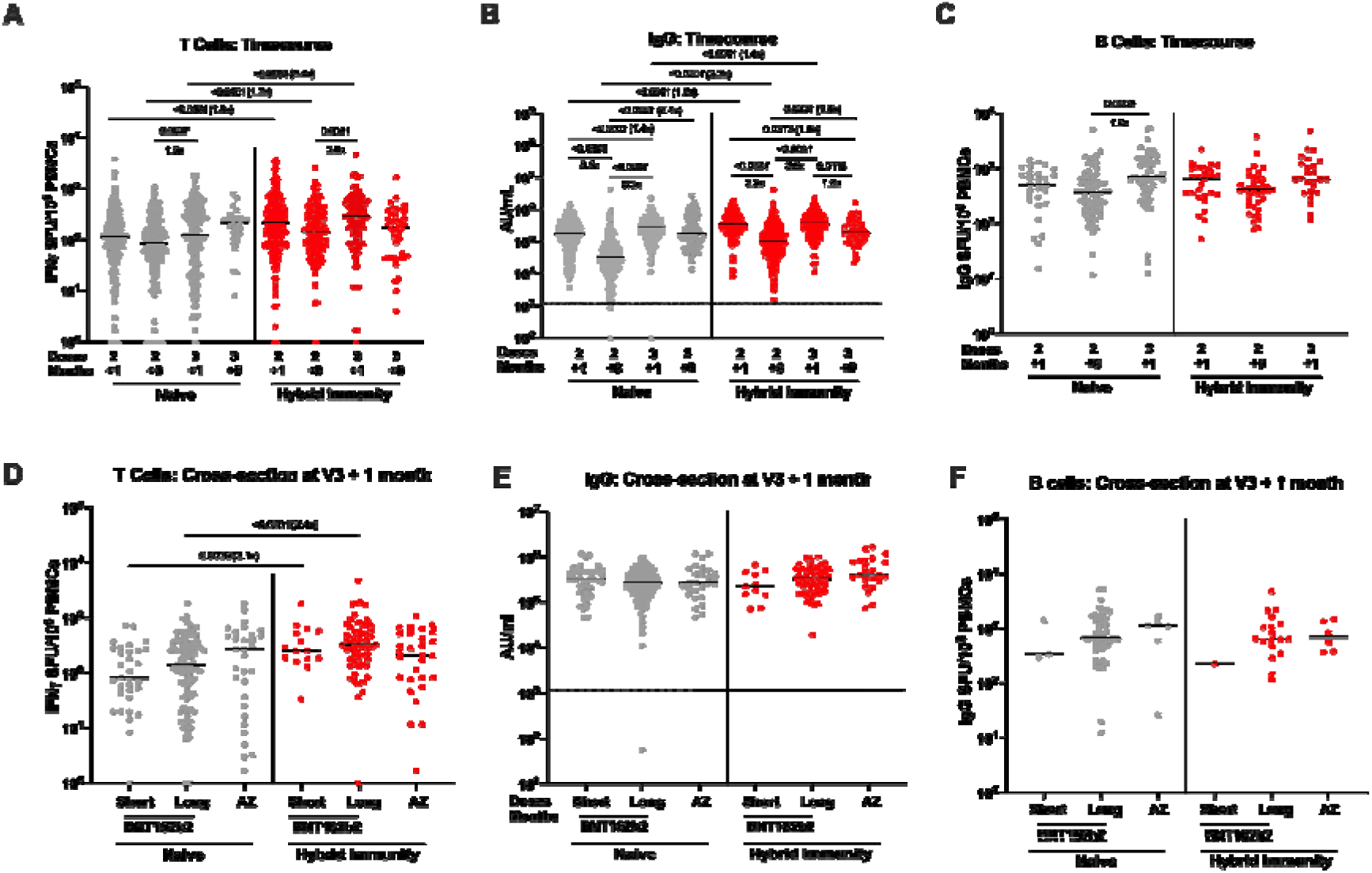
Time course of T cell, binding IgG and B cell responses for all participants, and cross section of responses one month post dose 3 after 2 doses of BNT162b2 (short or long interval) or AZD1222 vaccine. (A) Time course comparison of T cell responses to SARS-CoV-2 spike by IFNγ ELISpot assay for all vaccine regimens up to 6 months post third dose (n=613). (B) Time course comparison of IgG antibody response to SARS-CoV-2 spike by MesoScale Discovery assay for all vaccine regimens up to 6 months post third dose (n=680). (C) Time course comparison of B cell responses to SARS-CoV-2 spike by B cell ELISpot assay for all vaccine regimens up to one month post third dose. (D) Comparison of T cell responses one month after the third booster dose by primary vaccine regimen (BNT162b2 Short, Long or AZD1222). (E) Comparison of IgG antibody responses one month after the third booster dose by primary vaccine regimen. (F) Comparison of B cell responses one month after the third booster dose by primary vaccine regimen. Grey circles = naïve individuals, red circles = hybrid immunity. Bars represent the median. Comparisons within groups were tested with the Kruskal-Wallis nonparametric test and Dunn’s multiple comparisons correction, with 2-tailed p values shown above linking lines for significant differences with p≤0.05. Unpaired comparisons between naïve and hybrid immune time points were tested with the Mann Whitney test.

### Infection leads to boosting of IFNγ ELISpot T cell responses following all vaccine regimens

Spike-specific T cell responses were higher in those with hybrid immunity compared with infection naïve. This was the case for both BNT162b2 vaccinated groups (Supplementary Figure S3A and D) but was not the case in the AZD1222 group (Supplementary Figure S3G). T cell responses were still higher one month post dose 3 in those who had been previously infected with SARS-CoV-2 (Figure 3A). However, by 6 months post dose 3 the spike-specific T cell response in naïve and hybrid immune participants was equivalent (Figure 3A). M and N responses were higher in BNT162b2 vaccinated participants with hybrid immunity, but this difference was not seen in the AZD1222 group. Between one month post dose 2 and one month post dose 3, even in the group who did not seroconvert to N, we detected a rise in the T cell response to M and N in the BNT162b2 long interval naïve group (the largest group), which became significant one month after the third dose (Supplementary Figure S4D), and appeared to increase still further at the 6 month time point (although this was not significant). Given that T cell responses are more sustained than antibody responses, this presumably reflects people who became asymptomatically infected but whose subsequent samples were taken after waning of the N antibody response. We saw no such change in the AZD1222 group (Supplementary Figure S4G).

### Humoral responses wane quickly but are boosted by third dose vaccination

After the second vaccine dose, binding antibody responses decreased sharply. The median SARS-CoV-2 spike IgG titre (MSD) decreased 5.6-fold in naive vaccine recipients and 3.3-fold in the hybrid immunity group by 6 months (Figure 3B). Participants who received the different vaccine regimens followed similar patterns (Figure S3B, E and H). Naïve participants who received AZD1222 had lower spike antibody titres post second dose than those receiving BNT162b2 regimens, but these titres were then boosted 25-fold by the third (BNT162b2 mRNA) vaccine dose (Figure S3H). One month after the third dose, spike antibody IgG binding levels increased back to similar levels to those measured one month post dose 2 (Figure 3B). By 6 months after the third dose, the rate of waning was less than after the second dose, and was less in the naïve group than in the hybrid immune group. The naïve group waned by 1.4-fold between one and 6 months after the third dose, which was not significant, compared with 5.8-fold after the second dose. The hybrid immune group waned 1.9-fold between one and 6 months post dose 3, compared with 3.2-fold in the equivalent period after the second dose. The reduction was significant for the hybrid group, and brought it down to a level equivalent to that of the naïve group by 6 months post dose 3.

After the third dose, there was no significant difference in the magnitude of the spike binding IgG response between vaccine regimens (Figure 3E). Overall, a subtle (1.4-fold) but significant increase in spike IgG remained between previously infected and naïve participants one month after the third dose (Figure 3B). The IgG levels measured post dose 3 were significantly greater than those measured post dose 2 in naïve participants, but those with hybrid immunity did not derive additional benefit from the levels one month post dose 2, although there was a substantial boost over the 6 month post dose 2 level in this group. The RBD binding response followed the same pattern as the total spike response (Supplementary Figure S4B, E, H) and N antibody titres were unchanged by vaccination in the hybrid immune group (Supplementary Figure S4C, F, I). However, some of the naïve participants did show rises in N antibody between one and 6 months post third dose. This time period corresponded to the very large wave of omicron BA.1 in the UK, and likely represents subclinical infection in some of our participants.

Memory B cell responses were measured by IgG ELISpot in a subset of 106 participants (Figure 3C, Supplementary Figure S3C, F and I). Six months after the second dose, memory B cell frequencies were similar between naïve and hybrid immunity group, and these responses were preserved, with no statistically significant difference from one month post second dose. In the whole dataset, memory B cell responses were not impacted by the third vaccine dose, though there was a significant increase in the BNT162b2 long interval and AZD1222 groups in both naïve participants, (Supplementary Figure S3F and I), and in the BNT162b2 long interval participants with hybrid immunity (Supplementary Figure S3F). Unlike the T cell IFNγ response, where there was still an advantage in those previously infected, there was no increase in the memory B cell response in those previously infected in any group (Figure 3C and F).

These data indicate that although antibody levels decline between the second and third vaccine doses, T and B cell responses are well maintained across this period. Hybrid immunity conferred an advantage on the magnitude of the T cell and antibody response at all timepoints, including after the third vaccine dose, but did not for the B cell response. The third vaccine dose boosted immunity back to previous levels, or greater, with a tendency to even out any earlier differences between two-dose vaccine regimens.

Our cohort was mostly female, but the BNT162b2 short interval group contained significantly more male participants (30 of 84, 40.5%, p=0.006, Chi squared test, Table 1). We did not detect any significant differences in responses between the three vaccine regimens, but to ensure that there was no potential for the imbalance in male participants to influence this, we ran regression models to investigate the influence of age, sex, previous SARS-CoV-2 infection and vaccine regimen on log_10_ transformed spike-specific IgG and T cell responses (see Supplementary Table regression analysis). Multivariable models indicated that previous infection was independently associated with both IgG and T cell responses, but that male sex was inversely associated with T cell responses (Supplementary Tables 1A and E). Multivariable models were used to explore the effect of sex within each vaccine regimen group, for IgG and T cell responses. Sex had no effect on IgG responses in all three vaccine groups (Supplementary Tables 1B, C and D). Responses were negatively associated with age and associated with previous infection in the BNT162b2 long interval group. For T cell responses, previous infection was associated with T cell responses in the BNT162b2 groups, and male sex was negatively associated with T cell responses only in the AZD1222 group (Table 2C). Therefore, the male imbalance did not affect the measurement of responses in the BNT162b2 short interval group. Although we found evidence that T cell responses to a booster mRNA vaccine are weaker in men who have received a primary course of AZD1222, this must be viewed with caution as it is based on only 24 participants.

### Polyfunctional T cell responses are detectable six months after vaccination, with enhancement in individuals with hybrid immunity

T cell responses measured by intracellular cytokine staining (ICS) were lower at 6 months post second dose in AZD1222 vaccinated participants compared with BNT162b2 recipients (Figure 4A), in line with the ELISpot findings. T cell function was similar between the two BNT162b2 groups, and there was less IL-2 and TNF made by the AZD1222 group (Figure 4A). These differences evened out in the hybrid immunity group. CD8^+^ T cells made a substantial fraction of the IFNγ, at least half on average (Figure 4B), with a trend to more in the AZD1222 group, as known for chimpanzee adenovirus vectored vaccines (Barnes et al., 2012). Very little IL-2 was made by CD8^+^ T cells; the overwhelming majority of the IL-2 response came from CD4^+^ T cells on a per individual basis, irrespective of vaccination regimen (Figure 4C). All groups of participants made polyfunctional T cell responses, which we defined as IFNγ/IL-2/TNF triple-positive cells (Figure 4D). There were no differences between vaccine regimens in those with hybrid immunity, who uniformly had polyfunctional responses detectable.

**Figure 4.**
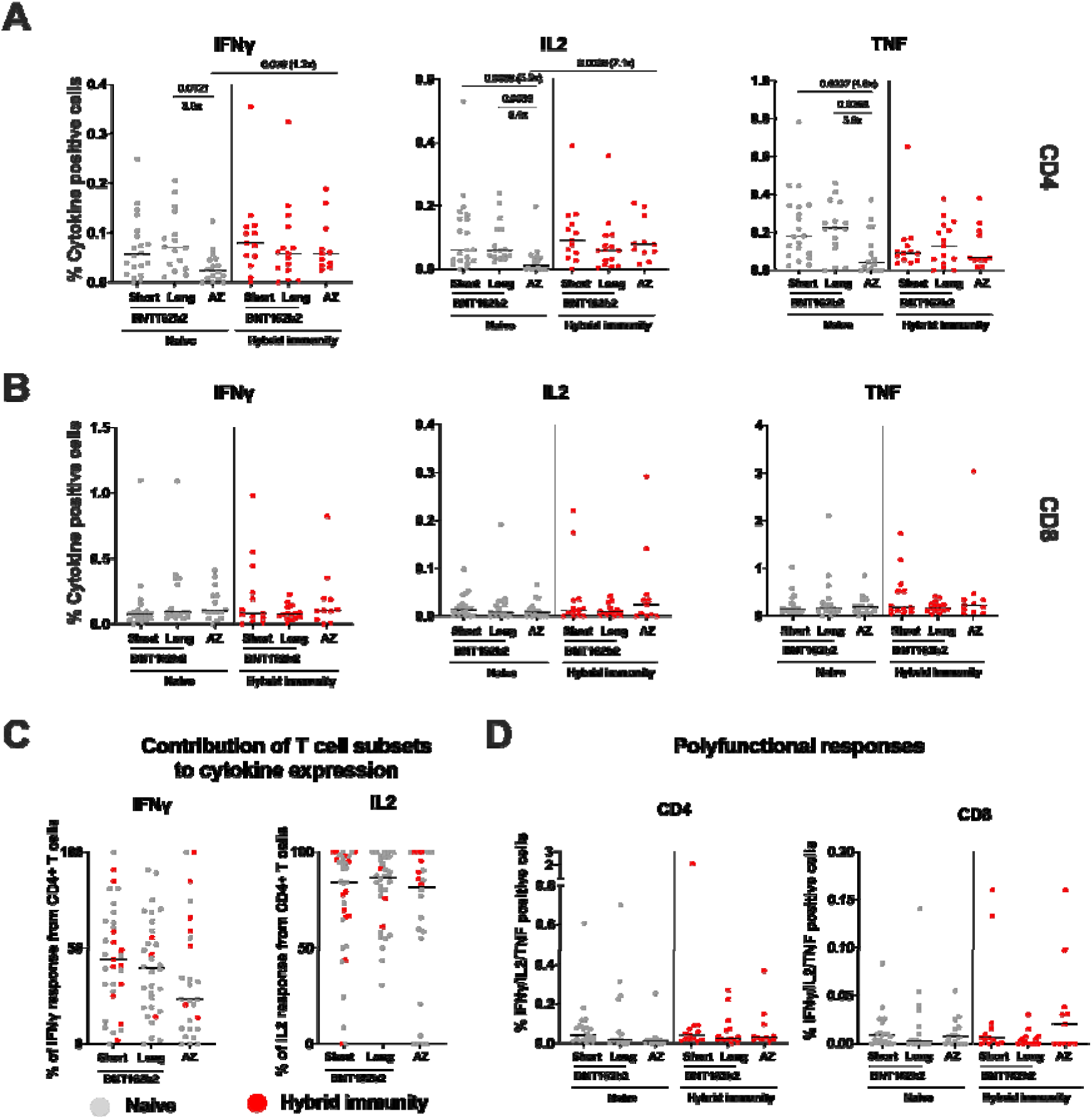
Analysis of spike-specific T cell responses by flow cytometry. Cryopreserved PBMCs from a subset of 95 participants who received BNT162b2 (Pfizer/BioNTech) with a short or long dosing interval, or AZD1222 (AstraZeneca), one month after the second dose, were analysed by intracellular cytokine staining and flow cytometry. The individual cytokine expression levels of total IFNγ, IL2 or TNF are shown as a percentage of (A) the CD4^+^ T cell population (top panels), or (B) the CD8^+^ T cell population (bottom panels). Populations were analysed by gating on single, live, CD3^+^ cells (Supplementary figure 1). Short = BNT162b2 short interval; Long = BNT162b2 long interval; AZ = AZD1222. Naïve participants are shown as grey circles and hybrid immunity group are red circles. Box plots represent the median, IQR and whiskers 1.5 x the IQR. (C) The T cell populations responsible for IFNγ or IL2 expression were assessed as the proportion of IFNγ or IL2 expressed by CD4^+^ T cells, calculated by dividing the cytokine production in CD4^+^ T cells by the total cytokine production in response to spike in both CD4^+^ and CD8^+^ T cells. (D) Polyfunctionality was evaluated by combined expression of IFNγ, IL2 and TNF in CD4^+^ and CD8^+^ T cells, showing the percentage of cells making all three cytokines. Naïve short: n=20, Naïve long: n=15, Naïve AZ n=14, Hybrid immunity short: n=13, Hybrid immunity long: n=17, Hybrid immunity AZ: n= 16. Unpaired comparisons across two groups were performed using the Mann Whitney test with 2-tailed p values shown above linking lines when 2-tailed p≤0.05.

### CD4+ and CD8+ proliferation responses to SARS-CoV2 spike are higher in previously infected participants

We also assessed cellular responses to SARS-CoV2 using T cell proliferation, a measure more biased towards central memory responses than IFNγ assays. T cell proliferation to spike S1 and S2 peptide pools was higher in previously infected AZD1222 vaccinated and the short interval BNT162b2 group compared to naïve individuals with a 3-8 fold increase in the median responses of CD4^+^ and CD8^+^ T cells respectively (Figure 5A, B and D), thus confirming the enduring increase in cellular memory conferred by infection combined with vaccination. As expected, responses to M and N were absent in the majority of naïve individuals (Figure 5C, E) with only one sample per vaccination regimen showing slightly elevated CD4^+^ T cell proliferation (3-11%) which was not explained by N seroconversion (Figure 5C). Differences between vaccination regimens were only apparent in the BNT162b2 vaccinated hybrid immunity groups with significantly increased CD8 responses to S1, S2 and M in the short compared to the long interval.

**Figure 5.**
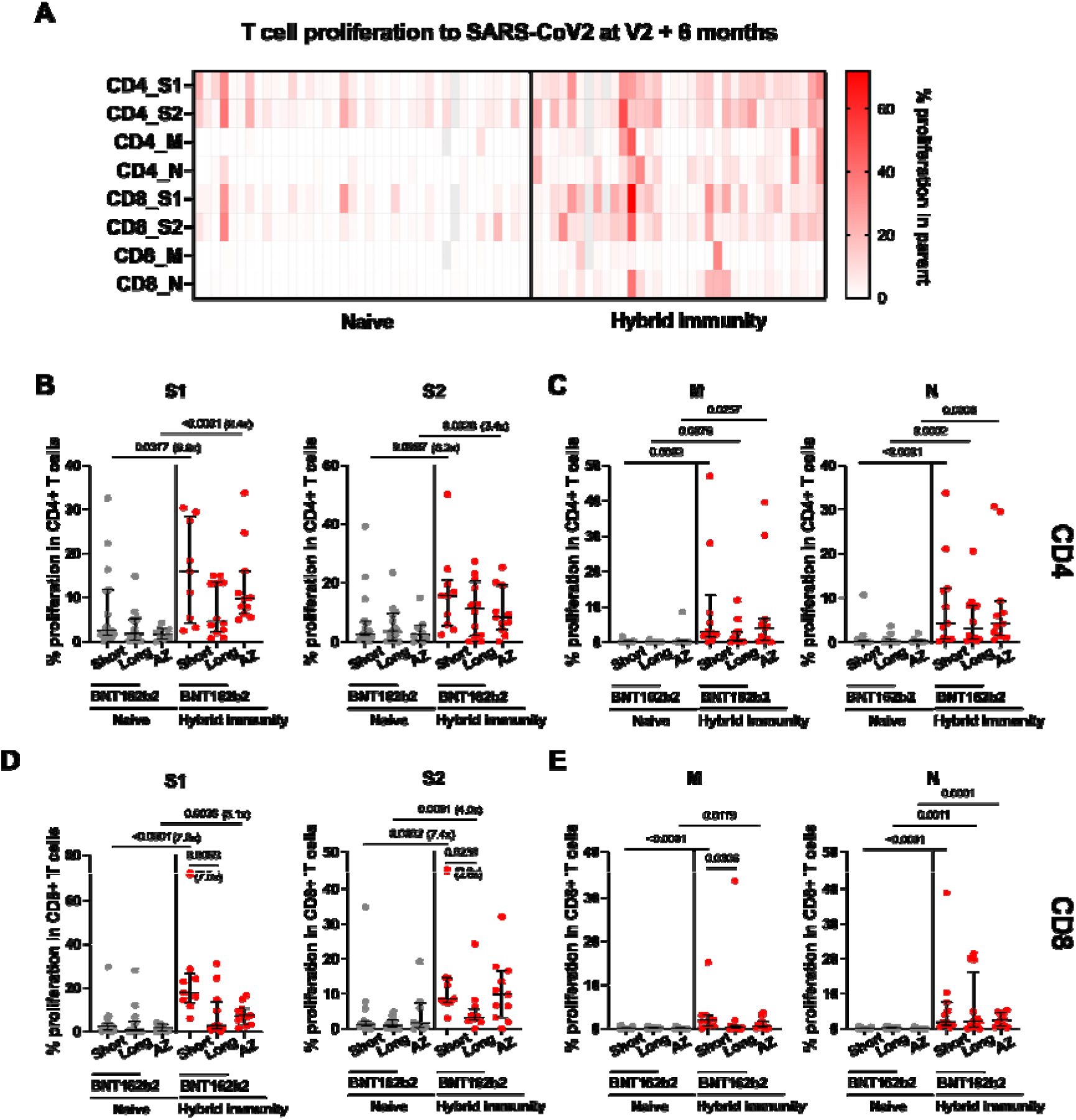
T cell proliferation to SARS-CoV2 at 6 months after the primary vaccine course of 2 doses of BNT162b2 or AZD1222. T cell proliferation to SARS-CoV2 peptide pools was assessed by flow cytometry in PBMC from 73 participants who had received either BNT162b2 with a short or long vaccine dosing interval or AZD1222 vaccine and were either naïve or were previously infected (either at baseline or during the course of the study). (A) Relative frequency of CD4^+^ and CD8^+^ T cells proliferating to individual peptide pools spike S1, spike S2, membrane (M) and nucleocapsid (N) protein in naïve (n=39) and hybrid immunity (n=34) individuals. Grey colour=missing value. (B, D) Proliferation to S1 and S2 and (C, E) M and N protein in CD4^+^ (B, C) and CD8^+^ (D, E) T cells are shown across the 3 vaccine regimens separated by exposure status (naïve versus hybrid immunity). Individual data points and median with IQR are displayed for naïve short: n=16, naïve long: n=15, naïve AZ: n=8, hybrid immunity short: n=11, hybrid immunity long: n=12, hybrid immunity AZ: n=11. Comparisons between naïve and hybrid immunity within each vaccine regimen were performed using the Mann Whitney test, and comparisons between the three vaccine regimens within the naïve and previously infected groups was performed using the Kruskal Wallis test and Dunn’s multiple comparisons correction. 2-tailed P values are shown only for statistically significant comparisons (p≤0.05). Fold change between medians of two groups are shown in brackets next to or under p value. (Fold change is not shown for those comparisons where there was no proliferation detected in one of the groups.) Grey circles = naïve individuals, red circles = participants with hybrid immunity.

### The antibody response to SARS-CoV-2 broadens after the third vaccine dose including enhanced neutralisation activity against omicron BA.1

Despite the differences between the naïve vaccine groups in binding antibody 6 months after the second dose (Figure 2B), there was no significant difference in neutralisation capacity of sera from these participants against the ancestral Victoria strain (Figure 6A). Neutralisation titres were lower against delta and lower still against omicron BA.1 compared with Victoria, as previously described (Dejnirattisai et al., 2022; Schmidt et al., 2021). The BNT162b2 long interval group had higher neutralising titres against delta than the short interval group, as they did 28 days after the second dose (Payne et al., 2021a). Using a surrogate neutralisation assay on the MSD platform, which measures inhibition of spike-ACE2 binding, we measured neutralisation of a wider range of variants. We also observed differences with the BNT162b2 long interval group having higher antibody titres than the other groups (Figure 6B). Although there was a trend for higher titres in the BNT162b2 short group compared to the AZD1222 group, this did not reach significance. The surrogate neutralisation assay showed a good correlation with the live virus focus reduction neutralisation assay for Victoria, delta and omicron variants (Supplementary Figure S5).

**Figure 6.**
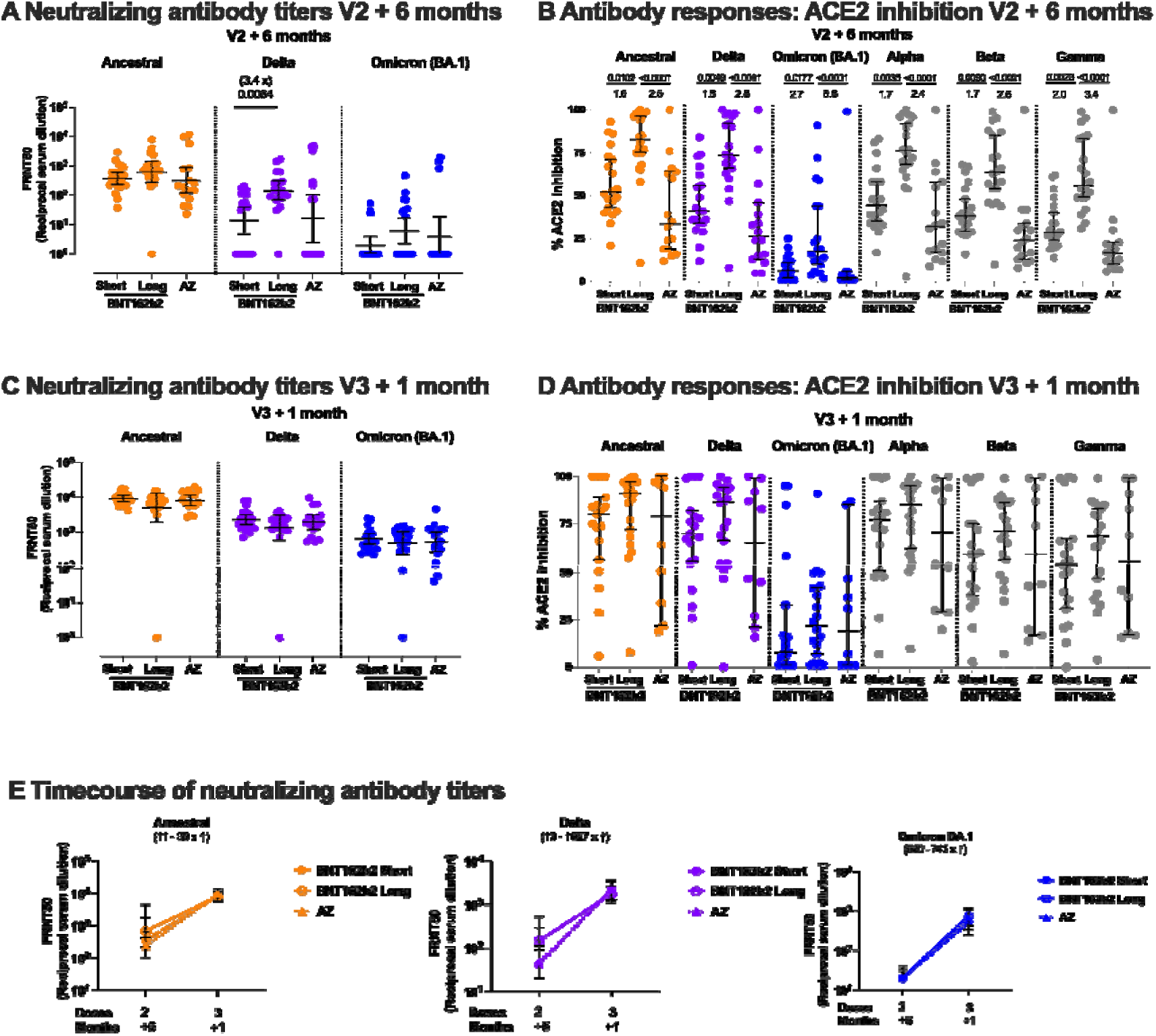
Neutralizing antibody and ACE2 inhibition titre profiles against SARS-CoV-2 variants of concern 6 months after 2 doses of BNT162b2 or AZD1222 and one month after a third vaccine with BNT162b2. Focus Reduction Neutralization Assay 50% (FRNT_50_) antibody titres against the Victoria isolate (orange), delta (B.1.617.2, purple) and omicron BA.1 (B.1.1.529 BA.1, blue) taken from infection-naïve participants. FRNT_50_ is the reciprocal dilution of the concentration of serum required to produce a 50% reduction in infectious focus forming units of virus in Vero cells (ATCC CCL-81). Participants either received 2 doses of BNT162b2 (Pfizer-BioNTech) vaccine delivered in a short (“Short”, 3-5 weeks, n=20) or long (“Long”, 6-17 weeks, n=20) dosing interval, or 2 doses of AZD1222 (AstraZeneca) vaccine (“AZ”, n=16). Neutralizing antibody titres are shown in (A) 6 months after the second dose, and (C) for the same individuals, one month after a third “booster” dose of mRNA vaccine for all participants. Geometric mean neutralizing titres with 95% confidence intervals are shown. (E) Comparison of the data from (A) and (C), plotted as means with error bars by vaccine regimen 6 months after the second vaccine (V2+ 6 months), one month after the third “booster” mRNA vaccine (V3+1 month). The range of fold change (median) between V2+6 months and V3+1 month for the three vaccine regimens (Short – dashed line, Long – solid line, and AZ – dotted line) is shown in brackets for each variant. Data in panels (A), (C) and (E) from the Short group (n=20) has been previously published (Dejnirattisai, Huo et al. 2022). (B) Impact of Short or Long BNT162b2 vaccine dosing interval and AZ on the ability of sera to inhibit ACE2 binding to SARS-CoV-2 spike (Victoria isolate, delta (B.1.617.2), Omicron BA.1 (B.1.1.529 BA.1), alpha (B.1.1.7), beta (B.1.351) and gamma (P.1) 6 months after the second dose and (D) one month after a third “booster” dose with mRNA vaccine. ACE2 inhibition was analysed using a multiplexed MSD® assay and performed at a serum dilution of 1:10 at V2 + 6 months and 1:100 at V3+1 months. Data are shown as percentage of inhibition. Bars represent the median with 95% confidence intervals. Naïve, Short: n=20; Naïve, Long: n=20; Naïve, AZ: n=16 for V2+6 months; Naïve, Short: n=19; Naïve, Long: n=20; Naïve, AZ: n=10 for V3+1 month. Vaccine regimens were compared with the Kruskal-Wallis nonparametric test and Dunn’s multiple comparisons correction, with 2-tailed p values shown above linking lines when 2-tailed p≤0.05, and fold changes are shown between the columns.

After the third dose of vaccine, neutralisation capacity against both the delta and omicron BA.1 variants increased. Our previous report in this cohort demonstrated that the neutralisation of omicron BA.1 was significantly higher 28 days after three doses of BNT162b2 compared to 28 days after 2 doses (Dejnirattisai et al., 2022). No differences were observed between vaccine groups after the third dose (Figure 6C). These differences also evened out in the ACE2 inhibition assay, though there was some saturation of the assay (Figure 6D). Therefore, although the overall level of binding antibody increased minimally (only in the naïve group) between 28 days after the second and 28 days after the third dose (Figure 3B), the neutralisation capacity of the antibody response broadened, and the gap between groups closed (Figure 6E). Thus, we observed a higher quality of response after the third dose, paralleling what has been seen for clinical effectiveness of a booster dose against omicron.

In a smaller subset of naïve participants, we extended these analyses to BA.2 and BA.3 (for MSD binding and ACE2 inhibition), 6 months post dose 2 and one month post dose 3. In order to determine the lasting effects of the booster dose on omicron variants post dose 3, we studied a further 115 participants for IgG binding to omicron variants, and 45 participants for live virus neutralisation to omicron BA.1, BA.2 and BA.5 6 months post dose 3. These assays showed that IgG binding to omicron BA.1, 2 and 3 spike was lower than that for the ancestral strain but persisted well 6 months after the third dose (Figure 7A-C), including binding to BA.4/5 which we measured at this time point. ACE2 inhibition by antibody was reduced for omicron BA.1-3, and ancestral and omicron responses waned (Figure 7D-F). However, the spread of responses at 6 months post dose 3 was wide, and by this point 11 of the 21 participants had contracted omicron infections (Figure 7F). Virus neutralisation for BA.1, BA.2 and BA.5 showed similar levels of neutralisation for BA.1 and BA.2, and a slight drop for BA.5 (Figure 7G). These responses waned significantly by 6 months, but in the subgroup of 11 people who became infected with between 1 and 6 months post dose 3 responses were significantly higher to omicron variants, but not to the ancestral virus (Figure 7G). Neutralisation responses correlated with ACE2 inhibition for most participants (Supplementary Figure S5D-I), with some evidence of saturation of the ACE2 assay. Importantly, overall, we detected less waning 6 months after the third dose than at the same time point after the second dose.

**Figure 7.**
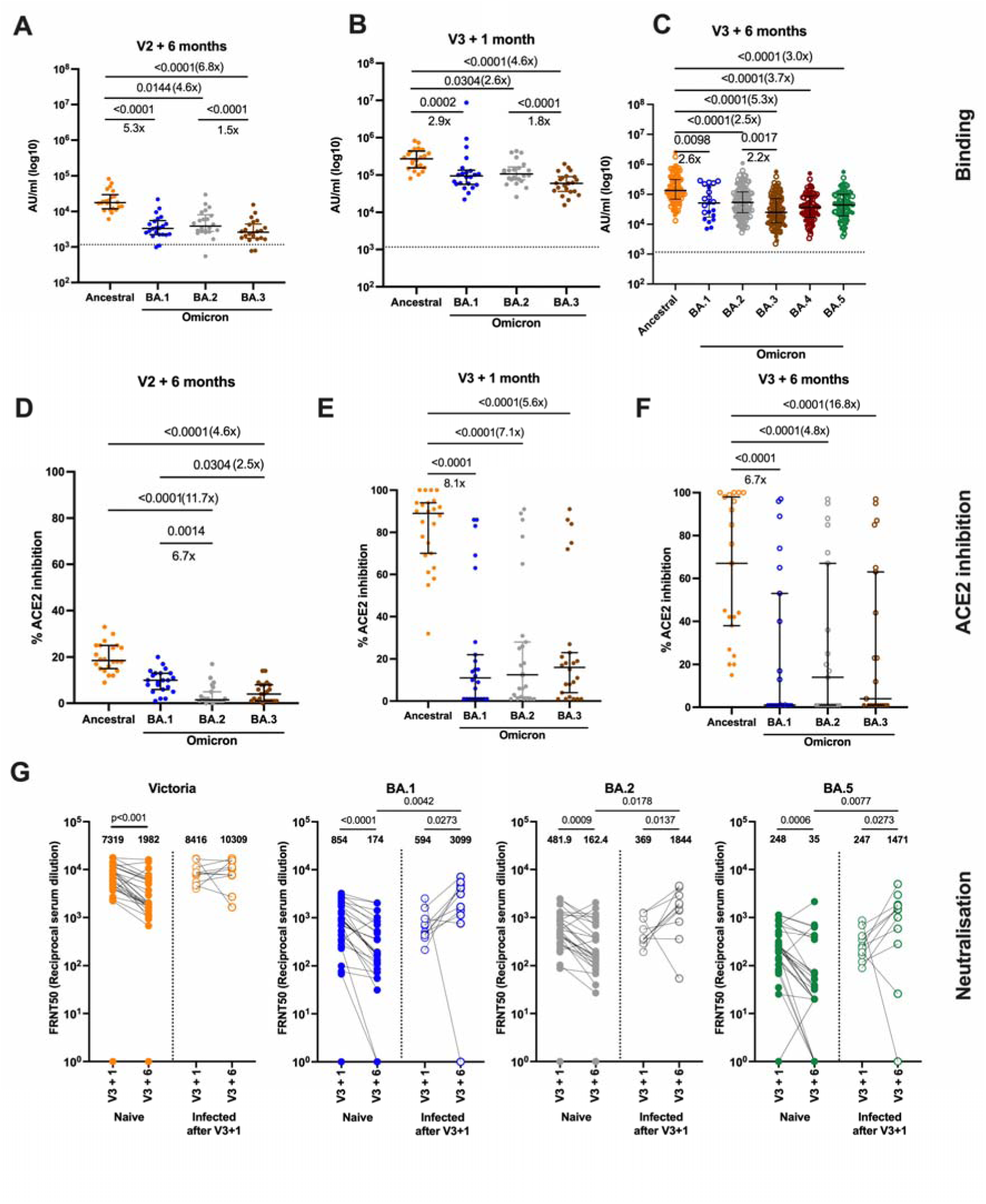
Antibody responses to omicron subvariants up to 6 months post dose 3. IgG binding measured on the MSD platform to spike from ancestral SARS-CoV-2 (orange) and the BA.1 (blue), BA.2 (grey), BA.3 (brown), BA.3 (maroon), BA.5 (green) omicron variants at 6 months after two doses of BNT162b2 (A) and one month post dose 3 of BNT162b2 vaccine in infection naïve participants (n = 21) (B) and 6 months post dose 3 of BNT162b2 vaccine (C) in both infection naïve participants (n=60) and in participants who became infected with an Omicron variant between 1 and 6 months post dose 3 (n=55). ACE2 inhibition by plasma from the same donors in A-C at 6 months post dose 2 (D), one month post dose 3 (E) and 6 months post dose 3 (F) ACE2 inhibition was performed at a serum dilution of 1 in 100 to account for saturation of the assay, as seen in figure 6. Comparisons between responses to ancestral and Omicron variants were made using Friedman’s test, with 2-tailed p-values of significant differences (p≤0.05) shown above linking line. (G) Neutralising antibody was measured at one month post dose 3 and 6 months post dose 3 by focus reduction neutralisation titre 50% (FRNT_50_) for Victoria strain (orange), BA.1 (blue), BA.2 (grey) and BA.5 (green) omicron variants in participants who remained infection naïve (n=33) and those who became infected in between one and 6 months post dose 3 (n=11). Filled circles indicate participants who remain infection naïve, participants who became infected with an omicron variant between one month and 6 months after the third vaccine dose are indicated in unfilled circles. Paired comparisons between one and 6 months post vaccine were tested using the Wilcoxon signed rank test, and comparis’n’s between groups were tested using the Mann Whitney test.

Cross reactive T and B cell responses to the omicron variant are preserved compared with the ancestral strain (Victoria) after second and third vaccine doses We investigated the effect of the third vaccine dose on T cell and B cell responses to omicron variants, in recognition of reduced vaccine effectiveness against infection with omicron but preservation of protection against severe disease. First, we tested responses to omicron BA.1 at 6 months post dose 2, similar to the situation for many people when omicron first appeared in the UK in November 2021. Unlike neutralising antibody responses, which were much lower for omicron BA.1 6 months after the second dose (Figure 6A), and lower but with the gap narrowed after the third dose (Figure 6C), T cell and B cell ELISpot responses were much less impacted. Using flow cytometry in the same participants in whom we studied multiple cytokine responses to spike, we did not detect any differences in the functionality of CD4 or CD8 T cell responses to omicron BA.1 at 6 months post dose 2 (Figure 8A and B), although the total proportion of the IFNγ response in CD4^+^ cells dropped slightly (Figure S6A).

**Figure 8.**
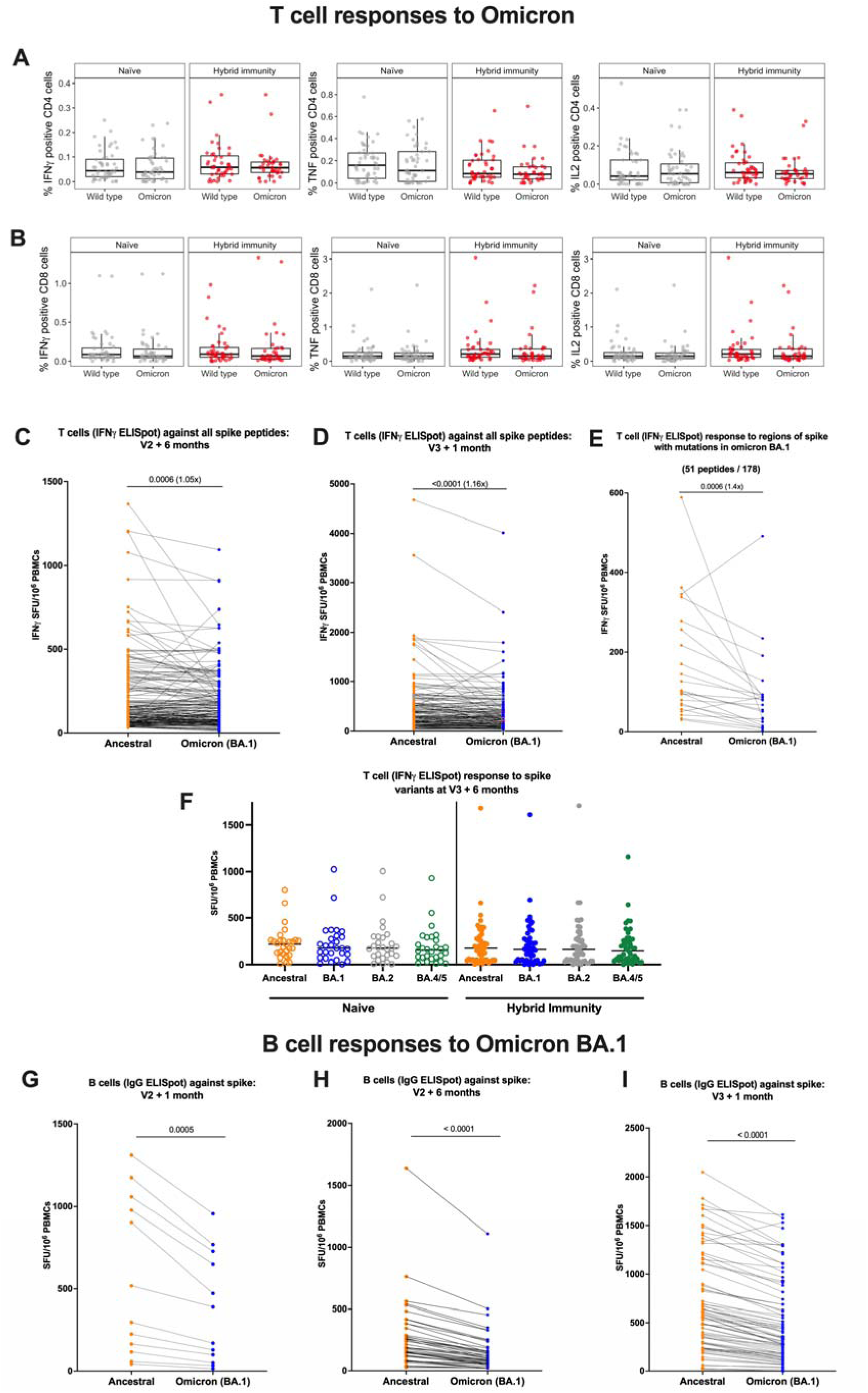
Comparison of cytokine response at 6 months post dose 2 against ancestral strain and omicron BA.1 variant according to infection status. Longitudinal comparison of T cell and B cell responses against ancestral strain and omicron BA.1 variant according to vaccine regimen and infection status. (A) Comparison of percentage IFNγ, TNF and IL2 positive CD4 T cells against ancestral strain and omicron BA.1 variant by intracellular cytokine staining of cryopreserved peripheral mononuclear cells (PBMCs) in either infection naive participants or participants with hybrid immunity. (B) Comparison of percentage IFNγ, TNF and IL2 positive CD8 T cells against ancestral strain and omicron BA.1 variant by intracellular cytokine staining of PBMCs in either infection naïve participants or participants with hybrid immunity. Pairwise comparison of T cell responses to spike from ancestral strain and omicron BA.1 variant from PBMCs by IFNγ ELISpot assay (C) in participants 6 months post primary vaccine course (2 doses of BNT162b2 or AstraZeneca), n=215, and (D) one month post third BNT162b2 vaccine dose, n=175. Displayed are responses to peptide pools representing the sum of S1 and S2 units of S from ancestral strain and omicron variant. (E) Pairwise comparison of IFNγ ELISpot responses in a subset of participants (n=36) to only the 51 out of 178 peptides spanning spike that have mutations in omicron BA.1 compared to the ancestral strain. (F) T cell responses to spike from ancestral strain and omicron BA.1, BA.2 and BA.4/5 variants in PBMCs from naïve (n=28) and hybrid immune (n=46) donors by IFNγ ELISpot assay. (G) Pairwise comparison of B cell responses to S in ancestral strain and omicron BA.1 variant from PBMCs in participants one month post vaccine dose 2 (n=12); (H) 6 months post second vaccine dose (n=43); (I) one month post third vaccine dose (n=80). Orange circles = responses against Victoria variant; blue circles = responses against omicron BA.1 variant. Displayed are responses to peptide pools representing S1 and S2 units of S from ancestral and omicron variants. ELISpot values are expressed as antibody SFU/10^6^ PBMCs. Horizontal lines represent median values. Comparisons between responses to ancestral and Omicron variants were made using Wilcoxon matched-pairs signed rank test, with 2-tailed p-values of significant differences (p≤0.05) shown above linking line.

Using the more sensitive IFNγ ELISpot assay, the proportion of ancestral SARS-CoV-2 T cell responses that were relatively preserved for omicron BA.1 on a per individual basis was very high 6 months after the second dose (median 94%, IQR 75-110), and one month after a third dose, (median 90%, IQR 70-104), although the difference between ancestral strain and omicron was significant by Wilcoxon matched pairs signed rank test (Figure 8C and D). Analysis of T cell ELISpot responses comparing only the peptides impacted by mutations did reveal a drop (Figure 8E, median 53%, IQR 22 -75), but this was not enough to have an impact on the T cell response for all of spike. We extended this analysis at 6 months post dose 3 for 46 hybrid immune and 28 naïve participants. We tested ancestral SARS-CoV-2 spike peptides alongside those from omicron BA.1, BA.2 and BA.4/5 (Figure 8F). At this point, there was no difference detected between the T cell response to any omicron variant in either group by 6 months post third vaccine dose.

For B cells, responses to omicron BA.1 were lower compared with the ancestral Victoria strain one month after the second dose (median 59% omicron relative to ancestral SARS-CoV-2, IQR 56-67, p=0.0005)(Figure 8G), 6 months after the second dose (median 57% IQR 45-64, p<0.0001)(Figure 8H) and one month after a third dose, (median 69% IQR 58-78, p<0.0001) (Figure 8I). This still represents a relative preservation of B cell immunity, compared with the absolute loss of neutralising antibodies to omicron after two vaccines (Figure 6A and C).

We also measured the effect of omicron on proliferative responses of T cells in some participants. No changes were observed for CD4^+^ and CD8 ^+^ T cell proliferation in the naïve group, though numbers of naïve participants were limited (Supplementary Figure S6B and D). In the hybrid immunity group, we observed a significant but modest drop in the proliferative response of CD4+ and CD8+ T cells to omicron BA.1 spike S2 (Figure S6C, p=0.0115) and S1 pool (Figure S6E, p=0.034) respectively when compared to ancestral spike. Overall, T and B cell responses to the omicron BA.1 variant were well preserved, compared with antibody responses.

## DISCUSSION

Our study reports robust immunity to SARS-CoV-2 spike including to omicron subvariants for all three primary vaccine regimens - BNT162b2 with a short (3-4 week) dosing interval, BNT162b2 with a long (6-17 week) dosing interval, and AZD1222 – following boosting with an mRNA vaccine. Over the course of the COVID-19 pandemic, vaccines have significantly reduced the link between the number of infections with SARS-CoV-2, and the numbers of hospital admissions and deaths due to COVID-19. Although there has been continual evolution of viral variants, which have evaded the antibody response to varying degrees (Harvey et al., 2021), vaccines have retained more effectiveness against severe disease than against overall infection (Andrews et al., 2022; Tartof et al., 2022; UK Health Security Agency, 2022). Emerging evidence implicates T cells as one potential mechanism for this protection, perhaps in addition to non-neutralising antibody functions (Bartsch Yannic et al.; Kaplonek et al., 2022b; Molodtsov et al., 2022; Scurr et al., 2022). The presence of both T cell and antibody responses gives the greatest protection from infection (Molodtsov et al., 2022) and from death in severe disease (Rydyznski Moderbacher et al., 2020), an observation that is also supported by studies in a macaque model (McMahan et al., 2021).

Here, in a cohort of participants which overlaps with the SIREN study - in which vaccine effectiveness has been shown (Hall et al., 2022), we have observed that responses after a third dose of COVID vaccine have different dynamics: binding and neutralising antibodies wane over the 6 months following the second dose, whereas B and T cell ELISpot responses wane much less over that interval. At 6 months post second dose, T cells secrete multiple cytokines and proliferate, indicating a broad range of memory function is retained by these cells. In addition, T cell responses are higher 6 months after vaccination in uninfected participants than they were in unvaccinated HCW 6 months after wave 1 infection in 2020, in a previous study of this cohort, (Tomic et al., 2022). Our findings are similar to those of Maringer et al. who also found that T cell responses were preserved more than antibody responses between the primary course and booster vaccination (Maringer et al., 2022), although we also found a benefit with the third dose, likely due to increased power from a much larger sample size.

The third vaccine dose boosted all responses from their nadir post dose 2. The relative magnitude of the T cell boost was smaller compared to the antibody boost but T cell responses had not waned to the same degree prior to the third dose. The third vaccine dose led to peak T cell levels which were higher than their previous peak one-month post second dose. In contrast, the boost to binding antibody response achieved by the third dose did not exceed the previous peak achieved post dose 2. Interestingly, although a third dose of vaccine did not achieve higher peak binding antibody levels, the neutralising capacity of the antibody response was much greater post dose 3 compared to post dose 2 – replicating earlier observations (Dejnirattisai et al., 2022). We observed that the B cell response also declined less in the 6 months after second vaccination than did the neutralising antibody response, and this implies many of these cells make antibody which binds, but does not neutralise, the virus.

With each successive vaccine dose, and up to one month after the third vaccine dose, participants who had been naturally infected with SARS-CoV-2 had their antibody and T cell responses boosted and the absolute values achieved were consistently higher than those who had not been naturally infected. These observations are particularly important when evaluating the relative benefit of a third vaccine dose which we demonstrate achieved statistically significant boosting effects even in the presence of hybrid immunity. These differences finally evened out by 6 months after the third dose. The ex-vivo immunogenicity benefits of hybrid immunity demonstrated here align with evidence of the enhanced clinical effectiveness of vaccination in the presence of hybrid immunity (Hall et al., 2022). Superior vaccine effectiveness has also been observed against omicron BA.1, BA.2 and BA.4/5 infections in those with hybrid immunity, compared with vaccination or infection alone (Altarawneh et al., 2022; Wei et al., 2022a). A recent systematic review comparing a range of estimates of protection from previous infection, vaccination and hybrid immunity has also found that hybrid immunity provides the greatest and most sustained protection (Bobrovitz et al., 2022).

We could also still detect an influence of the dose interval of BNT162b2 vaccine at 6 months after second vaccination. However, after the third vaccine dose, these differences had largely evened out and were no longer significant between the groups. T cell and antibody responses to spike were lower 6 months after primary vaccination course for AZD1222 compared to either BNT162b2 dosing regimen. These findings are compatible with previous reports for antibodies (Wall et al., 2021; Ward et al., 2022) and lower vaccine effectiveness against infection (UK Health Security Agency, 2022), although vaccine effectiveness against hospitalisation has been well preserved. After the AZD1222-primed recipients received a heterologous boost with mRNA vaccine, robust and similar cellular and antibody immunity including against omicron BA.1 variant was seen for all three regimens studied. We detected a possible influence of male sex on reducing T cell responses to a third dose of mRNA vaccine in people who had received a primary course of AZD1222 vaccine. However, this finding was based on a small number of participants so must be viewed with caution. The larger parent SIREN study would have greater potential to answer this question definitively, though the public health relevance of this observation is diminishing over time.

The third dose gave a broad immune response which could recognise all the variants tested. This included neutralisation of the omicron BA.1, BA.2 and BA.5 lineages. The few participants who were followed out to 6 months post dose 3 and had an omicron infection (11 participants) increased their neutralising antibody responses to omicron and not to Victoria, providing no evidence of immune imprinting (or antigenic sin) as has been recently suggested to occur with omicron (Reynolds et al., 2022). More recent population level evidence from Denmark and the UK suggests that omicron BA.1 or BA.2 infection in combination with vaccination is more protective against omicron BA.5 than alpha or delta infection (Hansen et al., 2022; Wei et al., 2022a). This may be due to waning immunity, antigenic difference, or both, rather than imprinting. We have not tested the effect of hybrid immunity on subsequent responses to omicron; such work is ongoing. However, we found no evidence of antigenic sin for responses after omicron infections, which were larger than the corresponding increase in antibody to the ancestral vaccine virus. T cell responses were less impacted by viral variants that antibodies, likely due to the wider range of epitopes available to T cells compared with antibodies, where protective responses are more focussed. Our findings are in line with those of others, who have also observed that antibodies decline more rapidly than T cell responses (Zhang et al., 2022). We found that T cell responses after the third dose were durable out to 6 months post dose, and that at this point, overall, ancestral and omicron strains were recognised equally well.

Our study has a number of limitations. (i) As with other HCW studies, our cohort has a female majority and is predominantly in people reporting white ethnicity. We have not observed any impact of sex or ethnicity in this study or our previous reports (Angyal et al., 2021; Payne et al., 2021a). (ii) Our longitudinal cohort does not include never-vaccinated participants, because all the HCWs engaged with our studies across six sites took up vaccination. However, we have been able to compare responses 6 months after vaccination (in 2021) with historical data using the same assay in a subset of the same cohort in 2020, 6 months after wave 1 (ancestral strain) infection before vaccine were available (Tomic et al., 2022) and demonstrate that vaccine-induced responses in infection-naïve HCWs are higher than infection-induced responses. (iii) We were not able to perform all assays on all participants at all timepoints, due to lack of sample availability, missed follow up visits, and/or laboratory capacity. This means that not all our data are longitudinal, though many are. To account for this, we have used unpaired testing in all our comparisons. (iv) We only performed neutralising antibody measurements on naïve participants due to the labour intensity and interpretation requiring matching with infecting variant strain and this information was limited. (v) We defined hybrid immunity in participants as previously testing PCR positive for SARS-CoV-2, or seroconversion to anti-N positivity during the study. However, some of the group labelled as naïve could have been exposed to SARS-CoV-2, because up to 60% of vaccinated people may not develop anti-N antibody, and the N sequence differs between variants (Follmann et al., 2022; Whitaker et al., 2021). As time went on, the N antibody levels rose in our naïve participants, even though many remained below the assay threshold for a positive N response. As hybrid immunity evolves in the population it will become increasingly difficult to define the shrinking group of people who have never been infected with SARS-CoV-2. (vi) For people with vaccine breakthrough infections since the second vaccine dose, infecting sequence data was not always available. However, we know that the majority of this report covers a period in time when delta was the predominant variant, with 68% and 88% of the sampling complete for this study by 1^st^ December 2021 and 1^st^ January 2022 respectively. (vii) Finally, we have not addressed mucosal immunity in this report, this is the subject of ongoing work. Antibody can be readily detected in the mucosa post infection with SARS-CoV-2 (Fröberg et al., 2021). Cellular and antibody responses have been also detected in the mucosa after COVID vaccination (Sano et al., 2022; Ssemaganda et al., 2022), but at low levels and their role in protection remains unclear.

In summary, we have observed that SARS-CoV-2 specific cellular immune responses are better maintained compared to antibodies in the 6 months following the second dose of COVID-19 vaccine. The third dose of vaccine confers a measurable benefit to these responses irrespective of the primary course, including in people who have previously been infected (“hybrid immunity”), who therefore may also stand to benefit from a third dose. The third dose also induces better antibody recognition of SARS-CoV-2 variants, including omicron BA.1. Despite public concern about loss of immunity over time post infection and/or vaccines, we find ample evidence of strong and durable immunity and memory responses that are likely to sustain protection against severe COVID-19 long term. Further booster vaccinations are likely to be most beneficial for preventing severe disease in the clinically vulnerable, and may lead to a reduction in hospitalisation rates. People with immune compromise are now receiving fourth or even fifth vaccine doses in UK and other countries, and parallel studies of durability of immunity in such populations are needed. The role of further booster vaccines for HCWs requires onward longitudinal follow-up of this cohort and others, but prevention of infection in HCWs continues to be desirable to minimise infection-related absence, nosocomial transmission and risks of long COVID (Antonelli et al., 2022). Our findings allow establishment of the dynamics of the immune response post infection and vaccination in a healthy population of working age, which can then be used as a benchmark for evaluating immunity in vulnerable groups, and provides the first glimpse of evolving “hybrid immunity” driven by ongoing viral exposure in vaccinated populations.

## METHODS

### Study design and sample collection

In this prospective, observational, cohort study, participants were recruited into the PITCH study from across six centres (Birmingham, Cambridge, Liverpool, Newcastle, Oxford and Sheffield). Individuals consenting to participate were recruited by word of mouth, hospital e-mail communications and from hospital-based staff screening programmes for SARS-CoV-2, including HCWs enrolled in the national SIREN study at three sites (Liverpool, Newcastle and Sheffield). Eligible participants were adults aged 18 or over, and currently working as an HCW, including allied support and laboratory staff, or were volunteers linked to the hospital. The majority of participants were sampled for previous reports in this PITCH cohort (Angyal et al., 2021; Ogbe et al., 2021; Payne et al., 2021a; Skelly et al., 2021). Participants were sampled for the current study between 4 January 2021 and 15 February 2022, with the majority of the sampling complete before the omicron BA.1 variant emerged in the UK (68% of sampling was prior to December 2021 and 88% was prior to January 2022).

Participants had received one of three vaccine regimens: “Short” - two doses of BNT162b2 (Pfizer/BioNTech) administered with the manufacturer’s licenced dosing interval (median 24 days, IQR 21-27); “Long” - two doses of BNT162b2 (Pfizer/BioNTech) administered with an extended dosing interval (median 71 days, IQR 66-78); and “AZ” - two doses of AZD1222 (Oxford/AstraZeneca), administered a median 74 days (IQR 65-78) apart. All participants then received a third “booster” dose of BNT162b2, a median of 207 days, (IQR 191-233) days after the second dose, regardless of primary vaccine regimen. Participants underwent phlebotomy for assessment of immune responses one (median 28 days, IQR 26-32) and six (median 185 days, IQR 173-200) months after the second dose of vaccine, and one month after the third dose of vaccine (median 31 days, IQR 28-37). Clinical information including BNT162b2 and AZD1222 vaccination dates, date of any SARS-CoV-2 infection (either prior to vaccination or during the study) defined by a positive PCR test and/or detection of antibodies to spike or nucleocapsid protein, presence or absence of symptoms, time between symptom onset and sampling, age, sex and ethnicity of participant was recorded. Key information on demographics and vaccine dose intervals is shown in Table 1.

Participants were considered to be SARS-CoV-2 exposed if they had ever been PCR or lateral flow device positive for SARS-CoV-2, irrespective of symptoms. In addition, participants were considered exposed to SARS-CoV-2 if they seroconverted with N antibody on the mesoscale discovery (MSD) assay. N seroconversion was defined as an N antibody level over the cut-off threshold of 3874 previously defined using pre-pandemic samples (Payne et al., 2021b), and at least a 2-fold increase over the baseline value. Participants who did not meet any of these criteria were considered to be infection-naïve.

PITCH is a sub-study of the SIREN study, which was approved by the Berkshire Research Ethics Committee, Health Research 250 Authority (IRAS ID 284460, REC reference 20/SC/0230), with PITCH recognised as a sub-study on 2 December 2020. SIREN is registered with ISRCTN (Trial ID:252 ISRCTN11041050). Some participants were recruited under aligned study protocols. In Birmingham participants were recruited under the Determining the immune response to SARS-CoV-2 infection in convalescent health care workers (COCO) study (IRAS ID: 282525). In Liverpool some participants were recruited under the “Human immune responses to acute virus infections” Study (16/NW/0170), approved by North West - Liverpool Central Research Ethics Committee on 8 March 2016, and amended on 14th September 2020 and 4th May 2021. In Oxford, participants were recruited under the GI Biobank Study 16/YH/0247, approved by the research ethics committee (REC) at Yorkshire & The Humber - Sheffield Research Ethics Committee on 29 July 2016, which has been amended for this purpose on 8 June 2020. In Sheffield, participants were recruited under the Observational Biobanking study STHObs (18/YH/0441), which was amended for this study on 10 September 2020. We also included some participants from Cambridge from a study approved by the National Research Ethics Committee and Health Research Authority (East of England – Cambridge Research Ethics Committee (SCORPIO study, SARS-CoV-2 vaccination response in obesity amendment of ‘‘NIHR BioResource’’ 17/EE/0025).The study was conducted in compliance with all relevant ethical regulations for work with human participants, and according to the principles of the Declaration of Helsinki (2008) and the International Conference on Harmonization (ICH) Good Clinical Practice (GCP) guidelines. Written informed consent was obtained for all participants enrolled in the study.

Peripheral blood mononuclear cells (PBMCs), plasma and serum were separated and cryopreserved. Some of the immune response data from one month after the second dose has been previously reported (Payne et al., 2021a), as has some of the neutralising antibody data for HCWs receiving a short dosing interval for BNT162b2 (Dejnirattisai et al., 2022). The study size was selected because this number was feasible for the six clinical and laboratory sites to study, and consistent with our track record of significant findings at this scale.

### Meso Scale Discovery (MSD) IgG binding assay

IgG responses to SARS-CoV-2, SARS-CoV-1, MERS-CoV and seasonal coronaviruses were measured using a multiplexed MSD immunoassay: The V-PLEX COVID-19 Coronavirus Panel 3 (IgG) Kit (cat. no. K15399U) from Meso Scale Discovery, Rockville, MD USA. A MULTI-SPOT^®^ 96-well, 10 spot plate was coated with three SARS CoV-2 antigens (Spike (S), Receptor-Binding Domain (RBD), Nucleoprotein (N)), SARS-CoV-1 and MERS-CoV spike trimers, spike proteins from seasonal human coronaviruses, HCoV-OC43, HCoV-HKU1, HCoV-229E and HCoV-NL63, and bovine serum albumin (negative control). Antigens were spotted at 200−400 μg/mL (MSD^®^ Coronavirus Plate 3). Multiplex MSD assays were performed as per the manufacturer’s instructions. To measure IgG antibodies, 96-well plates were blocked with MSD Blocker A for 30 minutes. Following washing with washing buffer, samples diluted 1:1,000-30,000 in diluent buffer, MSD standard and undiluted internal MSD controls, were added to the wells. After 2-hour incubation and a washing step, detection antibody (MSD SULFO-TAG™ anti-human IgG antibody, 1/200) was added. Following washing, MSD GOLD™ read buffer B was added and plates were read using a MESO^®^ SECTOR S 600 reader. The standard curve was established by fitting the signals from the standard using a 4-parameter logistic model. Concentrations of samples were determined from the electrochemiluminescence signals by back-fitting to the standard curve and multiplying by the dilution factor. Concentrations are expressed in Arbitrary Units/ml (AU/ml). Cut-offs were determined for each SARS-CoV-2 antigen (S, RBD and N) based on the mean concentrations measured in 103 pre-pandemic sera + 3 Standard Deviations. Cut-offs were: S, 1160 AU/ml; RBD, 1169 AU/ml; and N, 3874 AU/ml.

### MSD ACE2 inhibition assay

The V-PLEX SARS-CoV-2 Panel 23 (ACE2) Kit, from MSD, Rockville, MD, a multiplexed MSD immunoassay, was also used to measure the ability of human sera to inhibit ACE2 binding to SARS-CoV-2 spike antigens including B (Victoria), B.1.1.7/alpha, B.1.351/beta P.1/gamma, B.1.617.2/delta or B.1.1.529; BA.1/omicron BA.1). A MULTI-SPOT 96-well, 10 spot plate was coated with SARS-CoV-2 spike antigens including these ones above-mentioned. Multiplex MSD Assays were performed as per manufacturer’s instructions. To measure ACE2 inhibition, 96-well plates were blocked with MSD Blocker for 30 minutes. Plates were then washed in MSD washing buffer, and samples were diluted 1:10 – 1:100 in diluent buffer. Neutralizing activity was determined by measuring the presence of antibodies able to block the binding of ACE2 to SARS-CoV-2 spike proteins from Victoria spike, B.1.1.7/alpha, B.1.617.2/delta, B.1.351/beta, P.1/gamma and B.1.1.529; BA.1/omicron BA.1 and was expressed as percentage of ACE2 inhibition in comparison to the blanks on the same plate. Furthermore, internal controls and the WHO SARS-CoV-2 Immunoglobulin international standard (NIBSC 20/136) were added to each plate. After a 1-hour incubation, recombinant human ACE2-SULFO-TAG was added to all wells. After a further 1-hour, plates were washed and MSD GOLD Read Buffer B was added, plates were then immediately read using a MESO SECTOR S 600 Reader.

### Focus Reduction Neutralisation Assay (FRNT)

The neutralisation potential of antibodies (Ab) was measured using a Focus Reduction Neutralisation Test (FRNT), where the reduction in the number of the infected foci is compared to a negative control well without antibody. Briefly, serially diluted Ab or plasma was mixed with SARS-CoV-2 strain Victoria or P.1 and incubated for 1 hr at 37C. The mixtures were then transferred to 96-well, cell culture-treated, flat-bottom microplates containing confluent Vero cell monolayers in duplicate and incubated for a further 2 hr followed by the addition of 1.5% semi-solid carboxymethyl cellulose (Sigma) overlay medium to each well to limit virus diffusion. A focus forming assay was then performed by staining Vero cells with human anti-nucleocapsid monoclonal Ab (mAb206) followed by peroxidase-conjugated goat anti-human IgG (A0170; Sigma). Finally, the foci (infected cells) approximately 100 per well in the absence of antibodies, were visualized by adding TrueBlue Peroxidase Substrate (Insight Biotechnology). Virus-infected cell foci were counted on the classic AID ELISpot reader using AID ELISpot software. The percentage of focus reduction was calculated and IC50 was determined using the probit program from the SPSS package. In order to reduce confounding arising from exposure to different SARS-CoV-2 variants, these experiments were conducted only on participants who were naive at the time of sampling 6-months post second vaccine dose, as defined by no history of positive PCR or lateral flow test for SARS-CoV-2, and no anti-N IgG seroconversion during the study.

### T cell interferon-gamma (IFNγ) ELISpot Assay

The PITCH ELISpot Standard Operating Procedure has been published previously (Angyal et al., 2021). Interferon-gamma (IFNγ) ELISpot assays were set up from cryopreserved PBMCs using the Human IFNγ ELISpot Basic kit (Mabtech 3420-2A). A single protocol was agreed across the centres as previously published (Angyal et al., 2021) and available on the PITCH website (http://www.pitch-study.org/). In brief, PBMCs were thawed and rested for 3-6 hours in R10 media: RPMI 1640 (Sigma) supplemented with 10% (v/v) Fetal Bovine Serum (Sigma), 2mM L-Glutamine (Sigma) and 1mM Penicillin/Streptomycin (Sigma) in a humidified incubator at 37^∘^C, 5% CO_2_, prior to stimulation with peptides. PBMCs were then plated in duplicate or triplicate at 200,000 cells/well in a MultiScreen-IP filter plate (Millipore, MAIPS4510) previously coated with capture antibody (clone 1-D1K) and blocked with R10. PBMCs were then stimulated with overlapping peptide pools (18-mers with 10 amino acid overlap, Mimotopes) representing the spike (S), Membrane (M) or nucleocapsid (N) SARS-CoV-2 proteins at a final concentration of 2 µg/ml for 16 to18 hours in a humidified incubator at 37^∘^C, 5% CO_2_. For selected individuals, pools representing spike protein of the Omicron (BA.1) variant were included. Pools consisting of CMV, EBV and influenza peptides at a final concentration of 2µg/ml (CEF; Proimmune) and concanavalin A or phytohemagglutinin L (PHA-L, Sigma) were used as positive controls. DMSO was used as the negative control at an equivalent concentration to the peptides. After the incubation period as well as all subsequent steps wells were washed with PBS/0.05% (v/v) Tween20 (Sigma). Wells were incubated with biotinylated detection antibody (clone 7-B6-1) followed by incubation with the ELISpot Basic kit streptavidin-ALP. Finally colour development was carried out using the 1-step NBT/BCIP substrate solution (Thermo Scientific) for 5 minutes at RT. Colour development was stopped by washing the wells with tap water. Air dried plates were scanned and analysed with either the AID Classic ELISpot reader (software version 8.0, Autoimmune Diagnostika GmbH, Germany) or the ImmunoSpot® S6 Alfa Analyser (Cellular Technology Limited LLC, Germany). Antigen-specific responses were quantified by subtracting the mean spots of the negative control wells from the test wells and the results were expressed as spot-forming units (SFU)/10^6^ PBMCs. Samples with a mean spot value greater than 50 spots in the negative control wells were excluded from the analysis.

For comparison of responses to omicron BA.1 we firstly compared responses to 178 peptides spanning all of spike (S1 and S2) for the ancestral (wild type) and the omicron BA.1 variant, then secondly, we compared responses to the 51 peptides representing the regions of spike with mutations in omicron BA.1, again comparing ancestral and omicron BA.1. To reduce the disproportionate impact of background noise, samples with a total response to ancestral spike of <33 SFU/10^6^ PBMCs were excluded from analysis, with this cut off threshold calculated as the mean + 2 standard deviations of the DMSO wells across all experiments in the study. The % of the T cell response to ancestral strain that was preserved against omicron BA.1 was calculated for each paired sample then expressed as the median and IQR for the group.

### Memory B cell Fluorospot assay

Cryopreserved PBMCs were thawed and cultured for 72 hours with polyclonal stimulation containing 1 μg/ml R848 and 10 ng/ml IL-2 from the Human memory B cell stimpack (Mabtech). Using the Human IgA/IgG FluoroSpotFLEX kit (Mabtech), stimulated PBMCs were then added at 2x10^5^ cells/well to fluorospot plates coated with 10 μg/ml Sars-CoV-2 spike glycoprotein diluted in PBS. Plates were incubated for 16 hours in a humidified incubator at 37^∘^C, 5% CO_2_ and developed according to the manufacturer’s instructions (Mabtech). Analysis was carried out with AID ELISpot software 8.0 (Autoimmun Diagnostika). All samples were tested in triplicates and response was measured as spike-specific spots per million PBMCs with PBS background subtracted.

### Intracellular cytokine stimulation assay

In a subset of donors (n=95), selected at random from all three vaccine regimens and previous SARS-CoV-2 infection, T cell responses were characterised further using intracellular cytokine staining (ICS) after stimulation with overlapping SARS-CoV2 peptide pools. In brief, cryopreserved PBMCs were thawed, rested for 4-5 hours in R10 media and then plated at 1x10^6^ cells/well in a 96 well U-bottom plate together with co-stimulatory molecules anti-CD28 and anti-CD49d (both BD). Peptide pools (spanning ancestral (B.1) spike, omicron BA.1 spike, ancestral membrane (M) and nucleocapsid (N) proteins) were added at 2 μg/ml final concentration for each peptide. DMSO (Sigma) was used as the negative control at the equivalent concentration to the peptides. As a positive control, cells were stimulated with 1x cell activation cocktail containing phorbol-12-myristate 13-acetate (PMA) at 81µM and ionomycin at 1.3µM final concentration (Biolegend). The cells were then incubated in a humidified incubator at 37°C, 5% CO_2_ for 1 hour before incubating for a further 15 hours in the presence of 5µg/ml Brefeldin A (Biolegend). Flow cytometry staining was performed as described below.

### Proliferation assay

T cell proliferation assessed the magnitude of memory responses to SARS-CoV2 spike, M and N protein in the CD4^+^ and CD8^+^ T cell pool in 73 individuals selected for the ICS assay, with 27 participants from the BNT162b2 short interval group (16 naïve and 11 with hybrid immunity), 27 participants from the BNT162b2 long interval group (15 naïve and 12 with hybrid immunity) and 19 participants from the AZD1222 group (8 naïve and 11 with hybrid immunity). CellTrace^TM^ Violet (CTV, Invitrogen) labelling and stimulation with SARS-CoV-2 peptide pools spanning ancestral spike (divided into two pools, S1 and S2), omicron (BA.1) spike (S1 and S2), ancestral M and N protein, as well as a control peptide mix, CEF (1μg/ml per peptide) was carried out as previously described (Ogbe et al., 2021). Cells were incubated in RPMI 1640 (Sigma) supplemented with 10% human AB serum (Sigma), 2mM L-glutamine (Sigma) and 1 mM Penicillin/Streptomycin (Sigma) in a 96 well U-bottom plate at 250,000 cells per well in single or duplicate depending on cell availability. DMSO added at the same concentration to SARS-CoV-2 peptides served as negative control and 2ug/ml PHA-L as positive control. Cells were placed in a humidified incubator at 37^∘^C, 5% CO_2_. Half a media change was performed on day 4 and cells were harvested for flow cytometry staining on day 7 as described below. Data were expressed as relative frequency of proliferating cells within single, live CD4+ T cells and CD8+ T cells respectively. Background was subtracted from stimulated samples and samples were excluded due to high background (DMSO control >2% proliferation in any T cell subset,) or less than 1000 events in the single, live CD3+ gate (10 samples in total were excluded). Responses to individual peptide pools and summed responses to total spike (S1+S2) and M+NP were reported.

### Flow cytometry straining and analysis

Details for antibodies are listed in Supplementary Table 2. All washes and extracellular staining steps for PBMC were carried out in cell staining buffer (Biolegend) for ICS samples and PBS for proliferation samples. At the end of the culture period, PBMCs were washed once and subsequently stained with near-infrared fixable live/dead stain (Invitrogen) together with a cocktail of fluorochrome-conjugated primary human-specific antibodies against CD4, CD8, CD14 (all Biolegend) as well as human Fc blocking reagent (Miltenyi Biotec) for ICS and CD3, CD4 and CD8 (all Biolegend) for proliferation samples. Cells were stained at 4°C in the dark for 20 minutes, followed by one wash. Proliferation samples were then fixed with a 4% formaldehyde solution (Sigma) for 10min at 4°C, washed and stored in PBS in the fridge for up to one day. ICS samples were fixed and permeabilized in Cytofix/Cytoperm buffer (BD) for 20 min at 4°C, washed with 1x Perm buffer (BD) once followed by staining with the following primary human-specific antibodies diluted in Perm buffer: CD3, IFN-γ, TNF (all Biolegend), IL-2 (eBioscience) for 20 min at 4°C followed by one wash in 1x Perm buffer. Cells were stored in cell staining buffer in the fridge for up to one day. Samples were acquired on a MACSQuant analyser 10 and X (Miltenyi Biotec) and analysis was performed using FlowJo software version 10.8.1 (BD Biosciences). Example gating strategies are shown in Supplementary Figure 1.

### Statistical analysis

Continuous variables are displayed with median and interquartile range (IQR). Unpaired comparisons across two groups were performed using the Mann-Whitney test, and across three groups using the Kruskal-Wallis test with Dunn’s multiple comparisons test. Paired comparisons were performed using the Wilcoxon matched pairs signed rank test. Two-tailed P values are displayed. Statistical analyses were done using R version 4.0.2 (R Foundation for Statistical Computing, Vienna, Austria. URL https://www.R-project.org/) using the tidyverse packages (Wickham et al., 2019) and GraphPad Prism 9.3.1.

## Data Availability

All data produced in the present study are available upon reasonable request to the authors

## ACKNOWLEDGEMENTS

We are grateful to all our healthcare worker colleagues who participated in the study. For the Birmingham participants, the study was carried out at the National Institute for Health Research (NIHR)/Wellcome Trust Birmingham Clinical Research Facility. Laboratory studies were undertaken by the Clinical Immunology Service, University of Birmingham.

## AUTHOR CONTRIBUTIONS

Conceptualization, L.T., S.J.D., P.K., T.dS., S.H., V.H., C.J.A.D., R.P.P., A.R., M.C., G.S.; Methodology, S.J.D., P.K., L.T., S.C.M., B.K., S.L., T.dS., C.J.A.D., A.R., M.C., G.S., C.D., N.G., S.H., V.H.; Formal Analysis, B.K., S.C.M., S.J.D., L.T., T.dS., C.D., S.L., D.T.S., W.D., A.S.D., S.A., J.D.W. Investigation, B.K., R.P.P., S.L., C.L., W.D., S.A., N.M., S.F., S.A-T., S.C.M., T.T., L.M.H., A.A., R.B., A.R.N., S.L.D., E.C.H., L.H.B., P.S., A.C., A.B-W., L.S.R., A.L., J.K.T., H.H., I.G., M.P., P.Z., T.A.H.N., J.M.N., P.A., E.P., T.M., I.N., A.H., A.Sh., L.S., D.G.W., A.B.; Resources, A.B., L.T., E.B.; Data Curation, S.C.M., A.D.; Writing – Original Draft, L.T., S.J.D., P.K., Writing – Review & Editing, B.K., S.C.M., S.L., T.dS., S.L.D., S.J., D.G.W., C.P.C., K.J., P.C.M., A.J.P., J.M., E.B., A.R., M.C., G.S.; Visualization, S.C.M., S.L., B.K., S.A., J.D.W., L.T., S.J.D., Supervision, B.K., C.P.C., K.J., J.F., A.J.P., S.L.R-J., J.E.D.T., R.P.P., J.M., E.B., S.H., V.H., C.D., C.J.A.D., A.R., M.C., G.S., T.dS., L.T., P.K., S.J.D., Project Administration, A.S.D., Funding Acquisition, P.K., S.J.D., L.T., T.dS., C.J.A.D., A.R., S.H., V.H.

## DECLARATION OF INTERESTS

This work was funded by the UK Department of Health and Social Care as part of the PITCH (Protective Immunity from T cells to Covid-19 in Health workers) Consortium, UKRI as part of “Investigation of proven vaccine breakthrough by SARS-CoV-2 variants in established UK healthcare worker cohorts: SIREN consortium & PITCH Plus Pathway” MR/W02067X/1, with contributions from UKRI/NIHR through the UK Coronavirus Immunology Consortium (UK-CIC), the Huo Family Foundation and The National Institute for Health Research (UKRIDHSC COVID-19 Rapid Response Rolling Call, Grant Reference Number COV19-RECPLAS).

E.B. and P.K. are NIHR Senior Investigators and P.K. is funded by WT109965MA. S.J.D. is funded by an NIHR Global Research Professorship (NIHR300791). T.dS is funded by a Wellcome Trust Intermediate Clinical Fellowship (110058/Z/15/Z). RPP is funded by a Career Re-entry Fellowship (204721/Z/16/Z). C.J.A.D. is funded by a Wellcome Clinical Research Career Development Fellowship (211153/Z/18/Z). J.M. and G.S. are funded by the Chinese Academy of Medical Sciences (CAMS) Innovation Fund for Medical Science (CIFMS), China (grant number: 2018-I2M-2-002), Schmidt Futures, the Red Avenue Foundation and the Oak Foundation. The Wellcome Centre for Human Genetics is supported by the Wellcome Trust (grant 090532/Z/09/Z). P.C.M. is funded by Wellcome (110110z/15/Z), the Francis Crick Institute, and the University College London Hospital NIHR Biomedical Research Centre. J.E.D.T. is supported by the Medical Research Council (MR/W020564/1) and (MC_UU_0025/12). L.T. is supported by the Wellcome Trust (grant number 205228/Z/16/Z), the National Institute for Health Research Health Protection Research Unit (NIHR HPRU) in Emerging and Zoonotic Infections (EZI) (NIHR200907) and the Centre of Excellence in Infectious Diseases Research (CEIDR) and the Alder Hey Charity. The HPRU-EZI at University of Liverpool is in partnership with UK Health Security Agency (UKHSA), in collaboration with Liverpool School of Tropical Medicine and the University of Oxford. D.G.W. is supported by an NIHR Advanced Fellowship in Liverpool. M.C., S.L., L.T., and T.T. are supported by U.S. Food and Drug Administration Medical Countermeasures Initiative contract 75F40120C00085. The Sheffield Teaching Hospitals Observational Study of Patients with Pulmonary Hypertension, Cardiovascular and other Respiratory Diseases (STH-ObS) was supported by the British Heart Foundation (PG/11/116/29288). The STH-ObS Chief Investigator Allan Laurie is supported by a British Heart Foundation Senior Basic Science Research fellowship (FS/18/52/33808). We gratefully acknowledge financial support from the UK Department of Health and Social Care via the Sheffield NIHR Clinical Research Facility award to the Sheffield Teaching Hospitals Foundation NHS Trust.

The views expressed are those of the author(s) and not necessarily those of the NHS, the NIHR, the Department of Health and Social Care or Public Health England or the US Food and Drug Administration.

S.J.D. is a Scientific Advisor to the Scottish Parliament on COVID-19 for which she receives a fee. A.J.P. is Chair of UK Dept. Health and Social Care’s (DHSC) Joint Committee on Vaccination & Immunisation (JCVI), but does not participate in policy decisions on COVID-19 vaccines. He was previously a member of the WHO’s SAGE. The views expressed in this article do not necessarily represent the views of DHSC, JCVI, or WHO. AJP is chief investigator on clinical trials of Oxford University’s COVID-19 vaccine funded by NIHR. Oxford University has entered a joint COVID-19 vaccine development partnership with AstraZeneca. G.S. sits on the GSK Vaccines Scientific Advisory Board and is a founder member of RQ Biotechnology.

---

**Figure S1.**
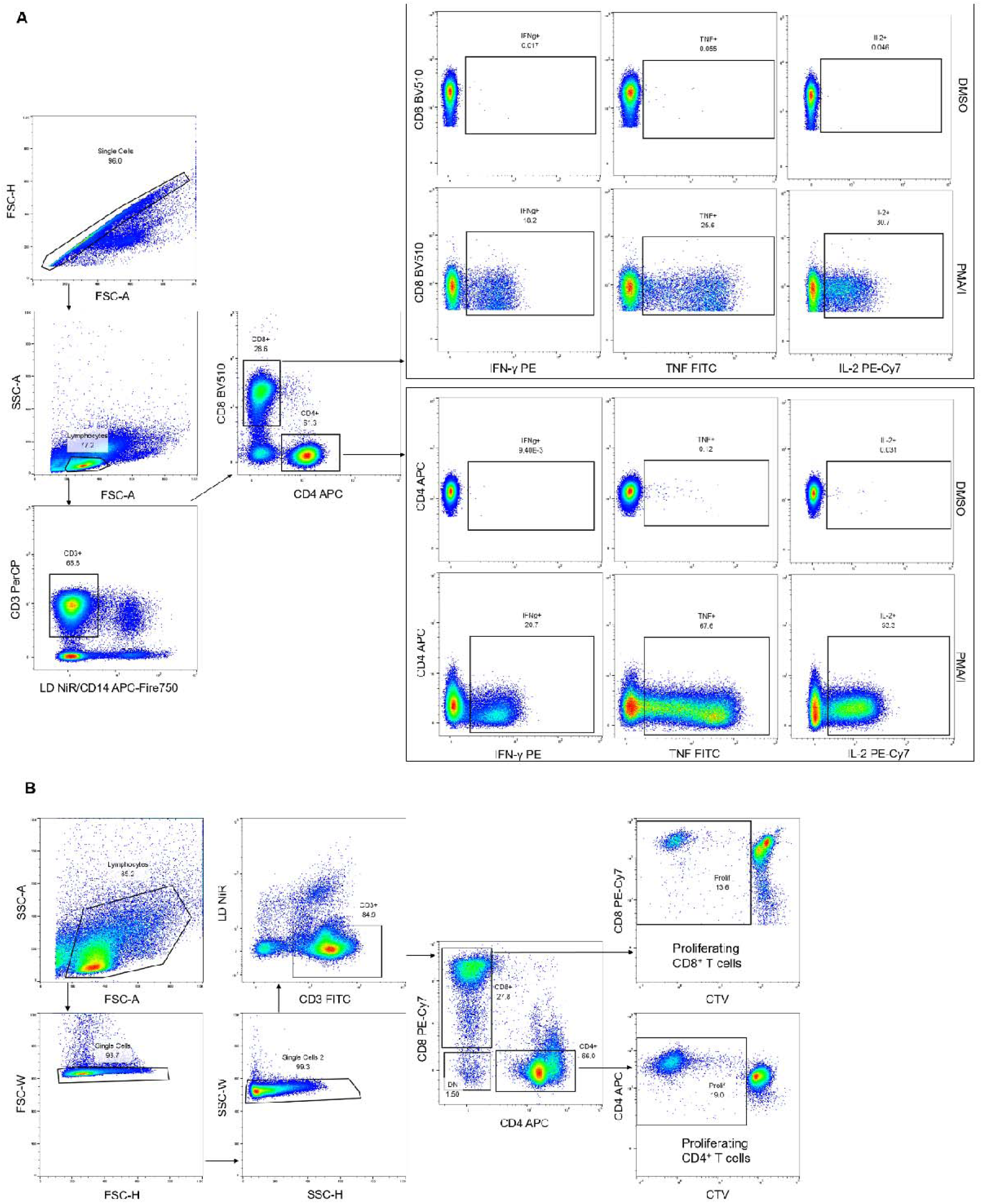
Gating strategy for T cell cytokine secretion (ICS) and proliferation. (A) For ICS assays single cells were gated using forward scatter (FSC)-area (A) and FSC-height (H) followed by a lymphocyte gate using FSC-A and side scatter (SSC)-A. Live CD3^+^ T cells were gated based on exclusion of dead cells (LD-NiR) and monocytes (CD14 APC Fire-750) as well as positivity for CD3 PerCP. T cell subsets were identified based on staining for CD4 APC and CD8 BV510 respectively and expression of cytokines (IFNγ, TNF, IL-2) was then identified in the CD4^+^CD8^-^ gate as well as the CD8^+^CD4^-^ gate. Representative gating is shown for the DMSO negative control and the PMA/Ionomycin positive control. In the case of PMA/Ionomycin the CD4^+^ gate was extended all the way to the CD4^-^ population due to downregulation of expression upon treatment (not shown in the figure). (B) For proliferation assays, lymphocytes were gated FSC-A and SSC-A parameters, followed by two subsequent single cell gates on FSC-H and width (W) as well as SSC-H and W to exclude doublets. From there live T cells were gated (LD-NiR low CD3^+^) and T cell subsets were identified (CD4^+^CD8^-^ and CD8^+^CD4^-^) using CD4^+^ APC and CD8^+^ PE-Cy7. Within the CD4^+^ and the CD8^+^ T cell gate proliferating cells were identified by gating on cells with reduced CTV (CellTrace^TM^ Violet) fluorescence intensity.

**Figure S2.**
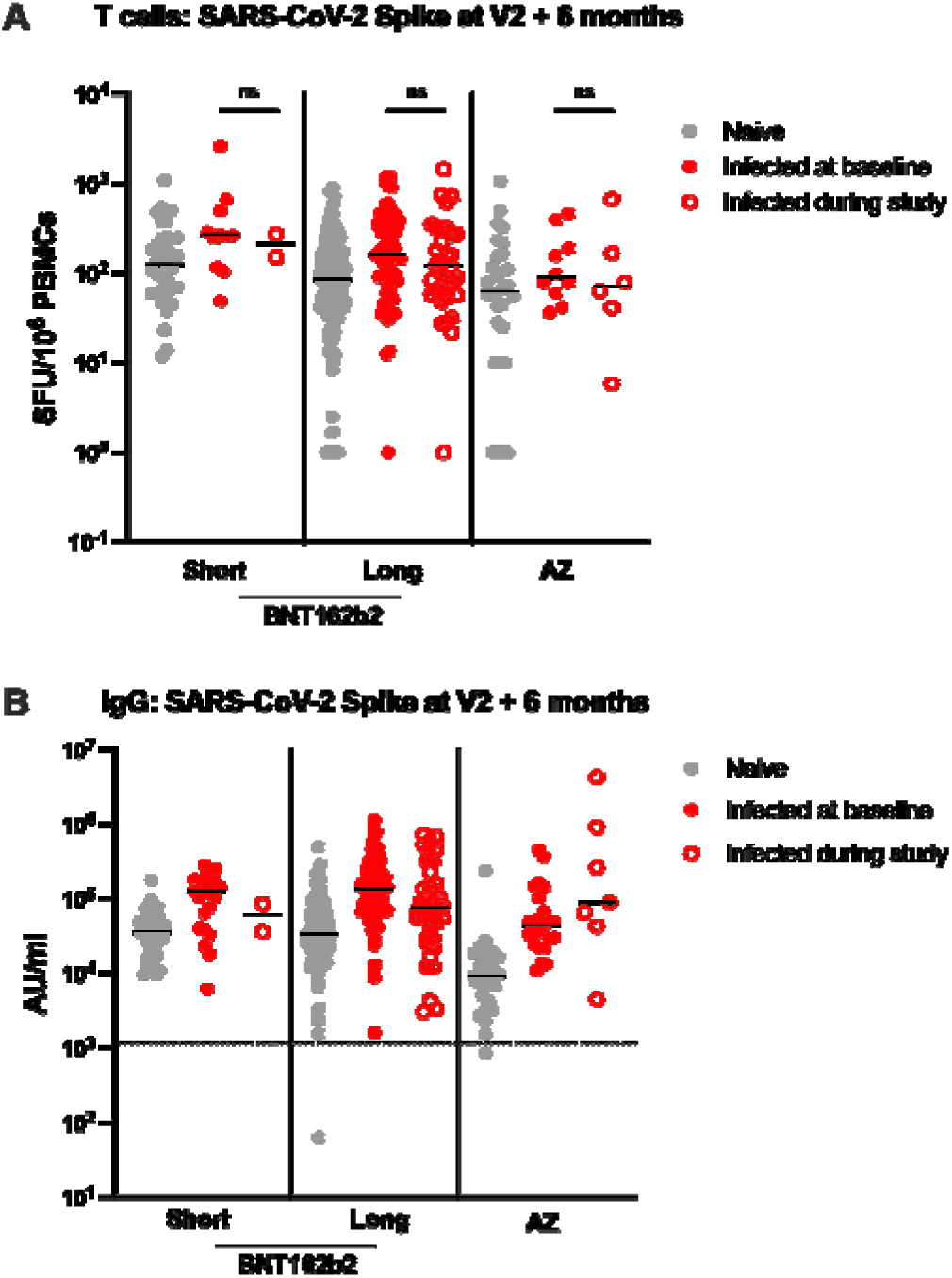
Comparison of T cell and IgG responses in those previously infected at baseline, infected during study, or infected at any time, at six months post second vaccine. (A) Comparison of IFNy ELISpot responses to S (ancestral strain) from cryopreserved PBMCs in short naïve (n=33), infected during study (n=2), previously infected at baseline (n=11) individuals; long naïve (n=116), infected during study (n=32), previously infected at baseline (n=62) individuals; AZ naïve (n=29), infected during study (n=6), previously infected at baseline (n=10) individuals. (B) Effect of vaccine regime and infection status on SARS-CoV-2 S-specific IgG responses in short naïve (n=38), infected during study (n=2), previously infected at baseline (n=21); long naïve (n=132), infected during study (n=36), previously infected at baseline (n=96); AZ naïve (n=27), infected during study (n=7), previously infected at baseline (n=23). Grey circles = naïve; solid red circles = previous infection at baseline; open red circles = infected during study. ELISpot values are expressed as SFU/10^6^ PBMCs, with values displayed responses to peptide pools representing S1 and S2 units of S (ancestral strain). IgG responses were measured in serum 6 months after the second dose using multiplexed MSD immunoassays and are shown in arbitrary units (AU)/mL. Horizontal bars represent the median. Vaccine regimens and vaccine status was compared using the Kruskal-Wallis test and Dunn’s multiple comparisons correction, with 2-tailed p-values shown above linking lines where significant (p≤0.05). ns = not significant.

**Figure S3.**
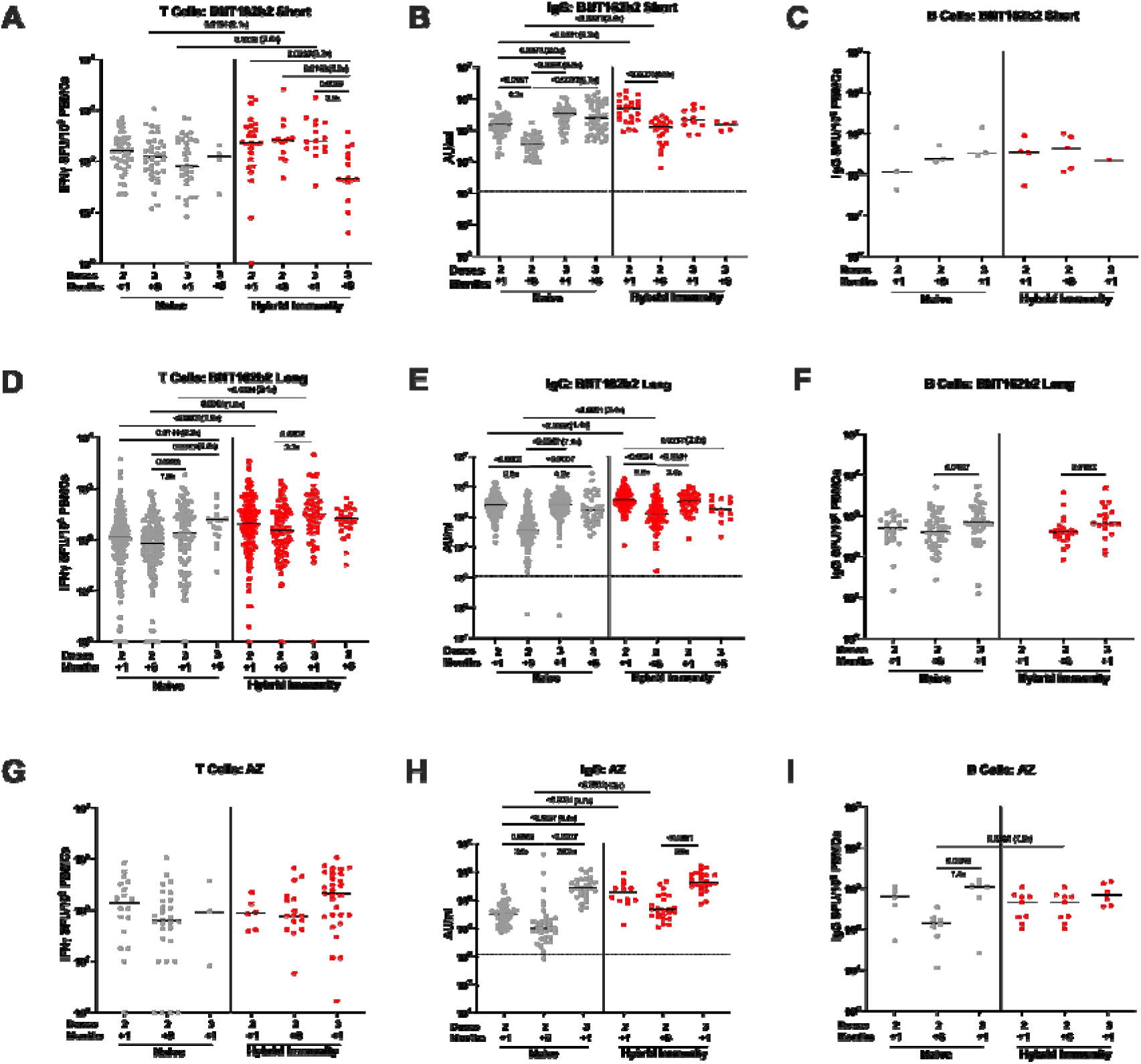
Time course of T cell, binding IgG and B cell responses 1 and 6 months after 2 doses of BNT162b2 (short or long interval) or AZD1222 vaccine: Responses are shown one and 6 months after 2 doses and following a third dose of BNT162b2 for (A) T cell responses to SARS-CoV-2 spike by IFNγ ELISpot assay after BNT162b2 (Pfizer-BioNTech) delivered with a short dosing interval (“Short”, 3-5 weeks, n=11-44 naïve, n=10-24 hybrid immunity), (B) IgG responses to SARS-CoV-2 spike by MesoScale Discovery (MSD) assay after BNT162b2 Short (n=24-59 naïve, n=8-24 hybrid immunity) and (C) B cell responses to SARS-CoV-2 spike by B cell Elispot assay after BNT162b2 Short (n=6-13 naïve, n=1-4 hybrid immunity). Responses are shown 1 and 6 months after 2 doses and following a third dose of BNT162b2 for (D) T cell responses to spike after a long interval (“Long”, 6-17 weeks, n=49-189 naïve, n=31-156 hybrid immunity), (E) IgG responses to spike by MSD assay after BNT162b2 Long (n=123-178 naïve, 78-203 hybrid immunity) and (F) B cell responses to spike after BNT162b2 Long (n=12-47 naïve, n=22-39 hybrid immunity). Responses are shown 1 and 6 months after 2 doses and following a third dose of BNT162b2 for (G) T cell responses to spike after AZD1222 (AstraZeneca) vaccine (“AZ”, n=18-26 naïve, 6-26 hybrid immunity), (H) IgG responses to spike by MSD assay after AZD1222 (n=28-54 naïve, n=16-44 hybrid immunity) and (I) B cell responses to spike after AZD1222 (n=5-8 naïve, n=7-10 hybrid immunity).

**Figure S4.**
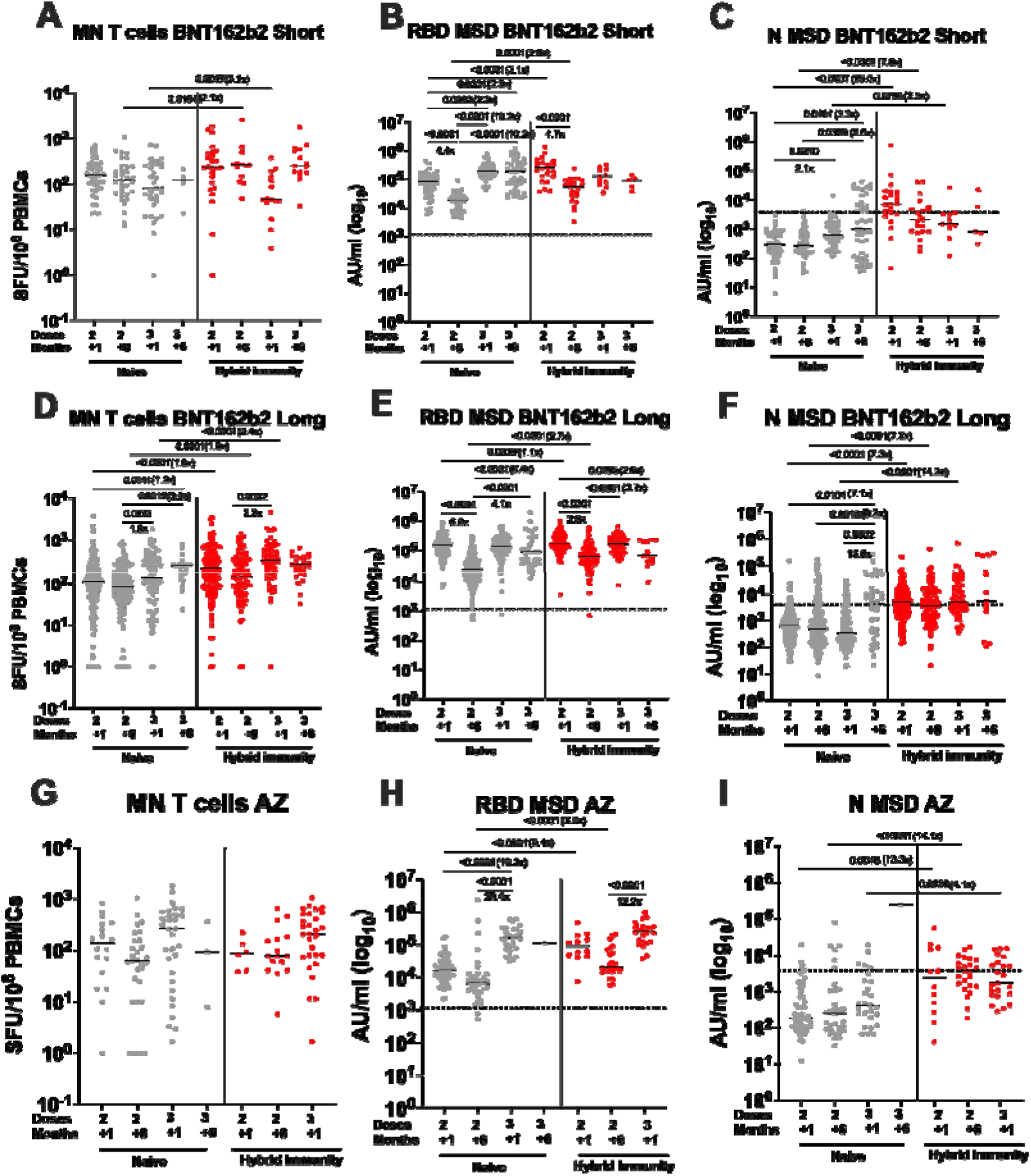
T cell and IgG Antibody responses to membrane protein, nucleocapsid protein and receptor binding domain. T cell and IgG antibody responses to membrane (M) and nucleocapsid (N) protein in participants receiving a primary course of BNT162b2 short dosing interval. (A) IFNy ELISpot responses in PBMCs, (B) IgG against receptor binding domain (RBD) and (B) IgG against nucleocapsid (N). T cell and IgG antibody responses to membrane (M) and nucleocapsid (N) protein in participants receiving a primary course of BNT162b2 long dosing interval. (D) IFNy ELISpot responses in PBMCs, (E) IgG against receptor binding domain (RBD) and (F) IgG against nucleocapsid (N). T cell and IgG antibody responses to membrane (M) and nucleocapsid (N) protein in participants receiving a primary course of AstraZeneca. (G) IFNy ELISpot responses in PBMCs, (H) IgG against receptor binding domain (RBD) and (I) IgG against nucleocapsid (N). Grey circles = naïve individuals, red circles = hybrid immunity. Bars represent the median. Comparisons are with the Kruskal-Wallis nonparametric test and Dunn’s multiple comparisons correction, with 2-tailed p values shown above linking lines for significant differences with p≤0.05.

**Figure S5.**
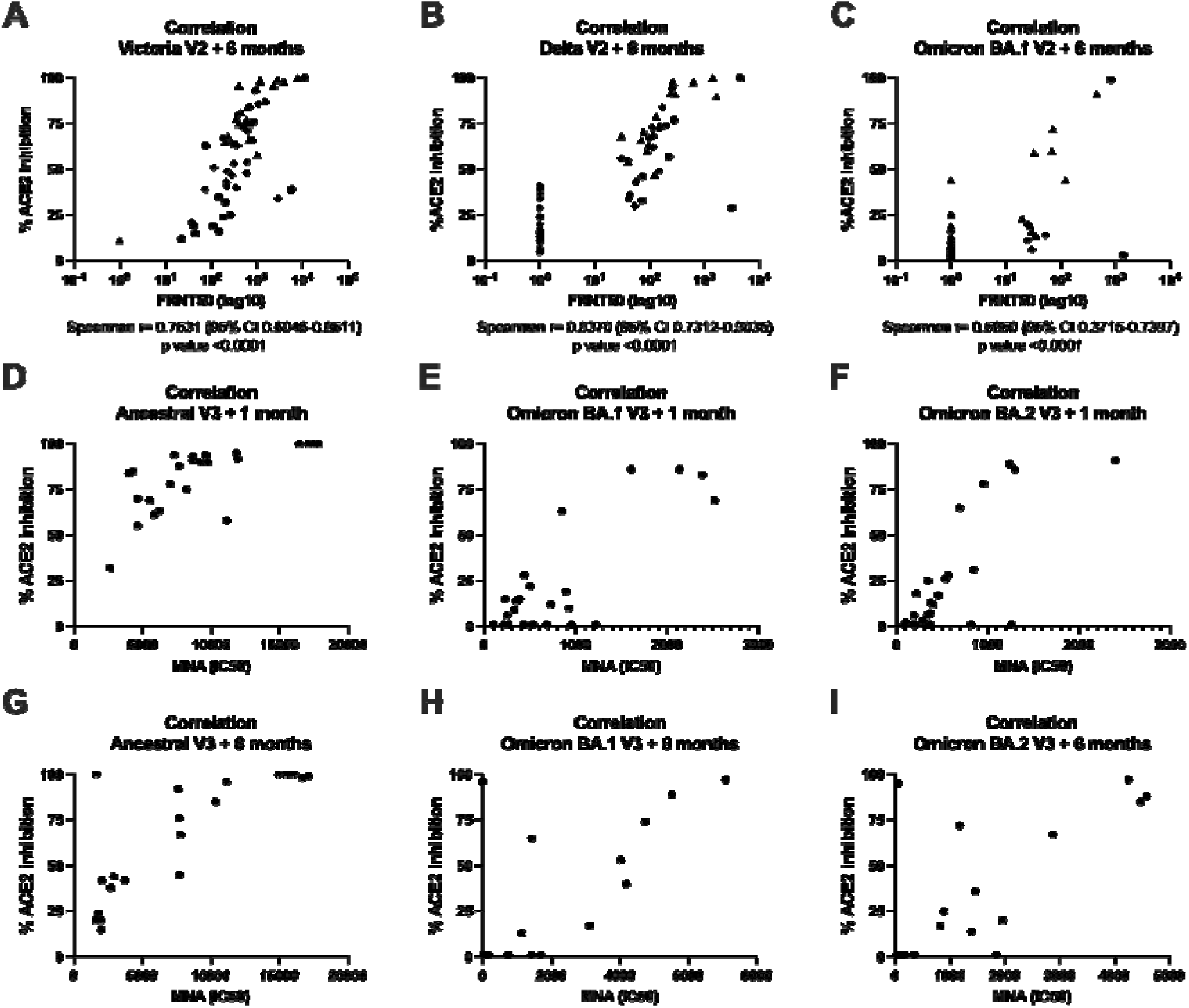
Correlation between ACE2 inhibition and neutralising antibodies. Correlation between the percentage of ACE2 inhibition and neutralisation titres against (A) Victoria, (B) Delta (B.1.617.2) and (C) Omicron BA.1 (B.1.1.529 BA.1), expressed as Focus Reduction Neutralization Assay 50% (FRNT_50_), determined in infection-naïve participants after receiving two doses of BNT162b2 (Pfizer-BioNTech) vaccine delivered in a short (“Short”, 3-5 weeks, n=20) or long (“Long”, 6-14 weeks, n=20) dosing interval, or two doses of AZD1222 (AstraZeneca) vaccine (“AZ”, n=15) 6 months after the second dose. Correlation between the percentage of ACE2 inhibition and neutralisation titres against (D) Ancestral, (E) Omicron BA.1 and (F) Omicron BA.2 expressed as half maximal inhibitory concentration (IC50), determined in infection-naïve participants one month after third vaccine dose. Correlation between the percentage of ACE2 inhibition and neutralisation titres against (G) Ancestral, (H) Omicron BA.1 and (I) Omicron BA.2 expressed as IC50, determined in infection-naïve participants 6 months after third vaccine dose. Pairwise correlations were assessed using Spearman’s rank-order correlation. Rhombus = Pfizer short, triangle= Pfizer Long, circle=AZ.

**Figure S6.**
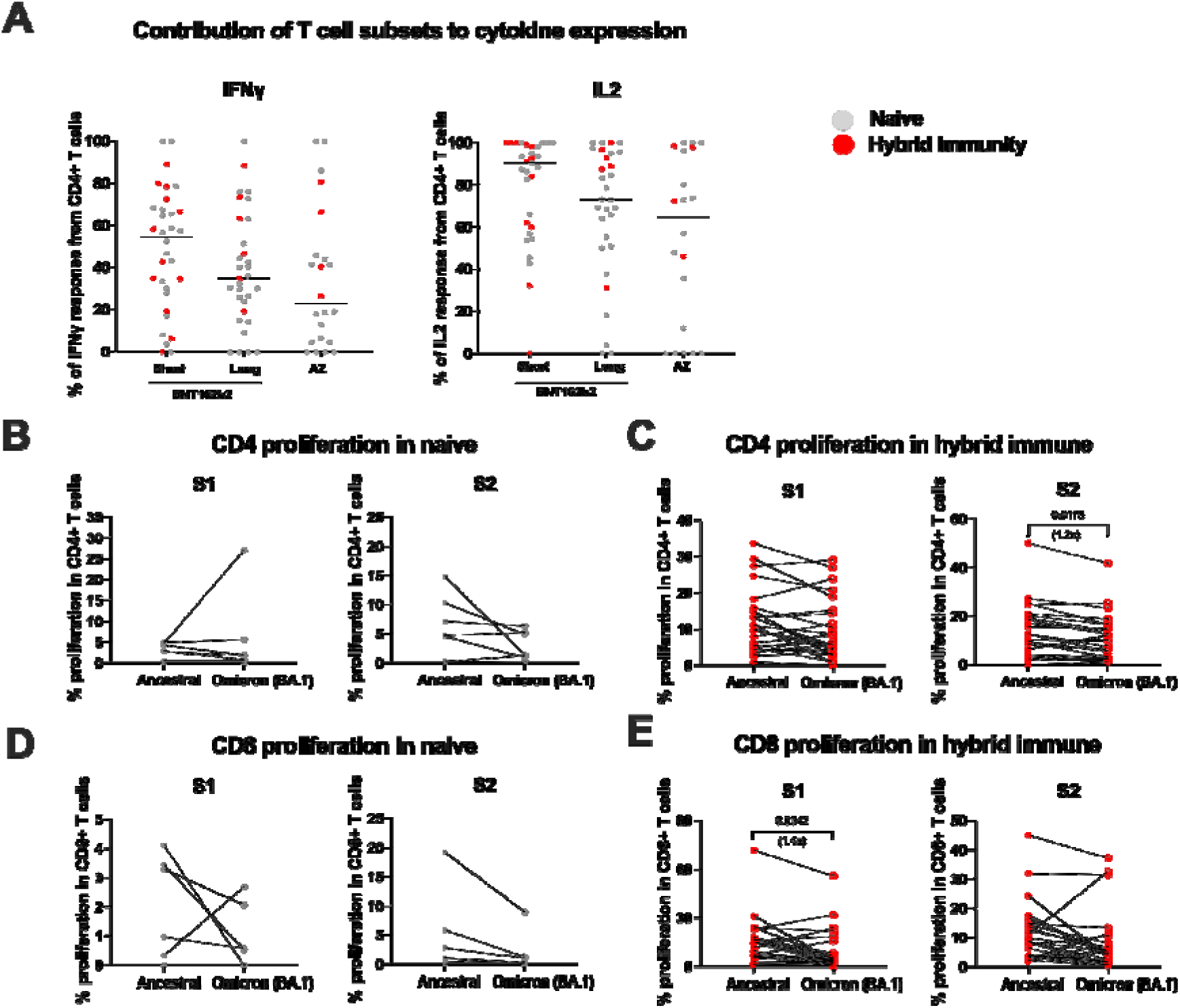
T cell cytokine responses and proliferation to the omicron (BA.1) variant 6 months after the primary vaccine course with BNT162b2 or AZD1222. (A) Combined data from naïve (grey) and hybrid immunity (red) participants shows cytokine responses (IFN-γ and IL-2) in CD4+ T cells at 6 months post second dose of either BNT162b2 (short and long dosing interval) or AZD1222. Proliferative responses of CD4^+^ and CD8^+^ T cells to SARS-CoV2 spike (S1+S2) from the ancestral strain were compared to the omicron BA.1 variant in a subset of (B, D) naïve (n=9) and (C, E) hybrid immunity (hybrid immunity, n=27) participants from all three vaccine regimens 6 months after the second dose. Individual data points are presented, and paired values are connected with a line. Paired testing was performed using Wilcoxon signed rank test and 2-tailed p values for significant differences (p≤0.05) are displayed. Closed circles = ancestral spike, open circles = omicron BA.1 spike, Grey = naïve individuals, red= individuals with hybrid immunity.

**Supplementary Table 2.**
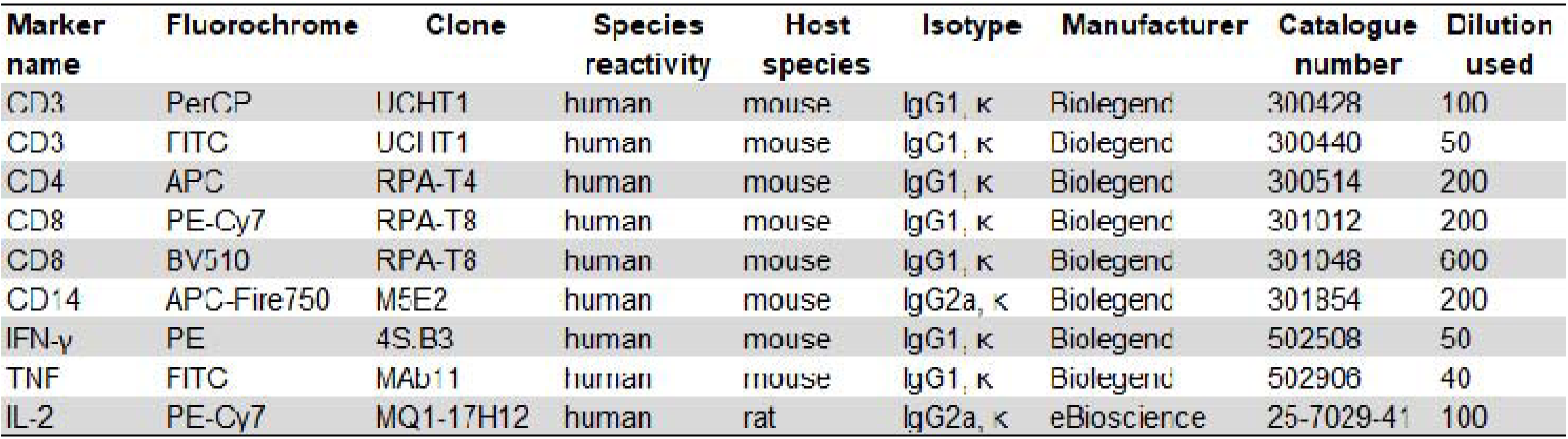
Antibodies used for intracellular cytokine staining and proliferation

## Notes

### Clinical Protocols

https://www.pitch-study.org

### Author Declarations

PITCH is a sub-study of the SIREN study, which was approved by the Berkshire Research Ethics Committee, Health Research 250 Authority (IRAS ID 284460, REC reference 20/SC/0230), with PITCH recognised as a sub-study on 2 December 2020. SIREN is registered with ISRCTN (Trial ID:252 ISRCTN11041050). Some participants were recruited under aligned study protocols. In Birmingham participants were recruited under the Determining the immune response to SARS-CoV-2 infection in convalescent health care workers (COCO) study (IRAS ID: 282525). In Liverpool some participants were recruited under the ‘Human immune responses to acute virus infections’ Study (16/NW/0170), approved by North West - Liverpool Central Research Ethics Committee on 8 March 2016, and amended on 14th September 2020 and 4th May 2021. In Oxford, participants were recruited under the GI Biobank Study 16/YH/0247, approved by the research ethics committee (REC) at Yorkshire & The Humber - Sheffield Research Ethics Committee on 29 July 2016, which has been amended for this purpose on 8 June 2020. In Sheffield, participants were recruited under the Observational Biobanking study STHObs (18/YH/0441), which was amended for this study on 10 September 2020. We also included some participants from Cambridge from a study approved by the National Research Ethics Committee and Health Research Authority (East of England — Cambridge Research Ethics Committee (SCORPIO study, SARS-CoV-2 vaccination response in obesity amendment of ‘NIHR BioResource’ 17/EE/0025). The study was conducted in compliance with all relevant ethical regulations for work with human participants, and according to the principles of the Declaration of Helsinki (2008) and the International Conference on Harmonization (ICH) Good Clinical Practice (GCP) guidelines. Written informed consent was obtained for all participants enrolled in the study.

### Summary of Updates

The below changes have been made following peer review: 1.We have added additional data on T cell ELISpot responses at 6 months after vaccine dose 3. These assays include measurement of responses against omicron BA.1, BA.2 and BA.4/5, alongside matching MSD binding data for the same participants at the same timepoint. 2.We have performed additional neutralising antibody assays at 1 month and 6 months post vaccine dose 3 on a further 19 participants nearly doubling the sample size for this assay. We have re-presented these data so that the participants who were infected between the third dose and the 6 month timepoint are shown separately from those that were not. 3.We have revised the section on ACE2 inhibition, and removed a potentially misleading phrase. 4.We have run linear regression models to examine the impact of sex as a confounder in the short/long interval BNT162b2 vaccinated participants. Sex imbalance in the BNT162b2 short interval group did not significantly influence the immune response overall in this cohort, the most important factor that affects the magnitude of the immune response is previous infection with SARSCoV2. We identified that men who received a primary course of AZD1222 vaccine had lower T cell responses, but this was based on a small number of participants and is of uncertain significance. 5.We have made various text changes as suggested by the reviewers, please see below for details on all these points. We have also added some more of the up to date literature. In addition, we have corrected some typographical errors that we identified in the submitted manuscript. 6.We have added an additional author who has helped respond to the reviewers comments by performing additional assays.

